# Cancer and the risk of COVID-19 diagnosis, hospitalisation, and death: a population-based multi-state cohort study including 4,618,377 adults in Catalonia, Spain

**DOI:** 10.1101/2021.05.18.21257371

**Authors:** Elena Roel, Andrea Pistillo, Martina Recalde, Sergio Fernández-Bertolín, María Aragón, Isabelle Soerjomataram, Mazda Jenab, Diana Puente, Daniel Prieto-Alhambra, Edward Burn, Talita Duarte-Salles

## Abstract

**Objectives:** To investigate the associations between cancer and risk of outpatient COVID-19 diagnosis, hospitalisation, and COVID-19-related death, overall and by years since cancer diagnosis (<1-year, 1-5-years, >5-years), sex, age, and cancer type.

**Design:** Population-based cohort study

**Setting:** Primary care electronic health records including ∼80% of the population in Catalonia, Spain, linked to hospital and mortality records between 1 March and 6 May 2020.

**Participants:** Individuals aged ≥18 years with at least one year of prior medical history available from the general population. Cancer was defined as any prior diagnosis of a primary invasive malignancy excluding non-melanoma skin cancer.

**Main outcome measures:** Cause-specific hazard ratios (aHR) with 95% confidence intervals for each outcome. Estimates were adjusted by age, sex, deprivation, smoking status, and comorbidities.

**Results:** We included 4,618,377 adults, of which 260,667 (5.6%) had a history of cancer. Patients with cancer were older and had more comorbidities than cancer-free patients. A total of 98,951 individuals (5.5% with cancer) were diagnosed and 6,355 (16.4% with cancer) were directly hospitalised (no prior diagnosis) with COVID-19. Of those diagnosed, 6,851 were subsequently hospitalised (10.7% with cancer) and 3,227 died without being hospitalised (18.5% with cancer). Among those hospitalised, 1,963 (22.5% with cancer) died. Cancer was associated with an increased risk of COVID-19 diagnosis (aHR: 1.08; 95% confidence interval [1.05-1.11]); direct COVID-19 hospitalisation (1.33 [1.24-1.43]); and death following a COVID-19 hospitalisation (1.12 [1.01-1.25]). These associations were stronger for patients recently diagnosed with cancer, aged <70 years, and with haematological cancers.

**Conclusions:** Patients recently diagnosed with cancer, aged <70 years, or with haematological cancers are a high-risk population for COVID-19 diagnosis and severity. These patients should be prioritised in COVID-19 vaccination campaigns and continued non-pharmaceutical interventions.

**What is already known on this subject:**

- Prior studies addressing the relationship between cancer and COVID-19 infection and adverse outcomes have found conflicting results
- The majority of these studies had small sample sizes, were not population-based (i.e. restricted to hospitalised patients), thus increasing the risks of selection and collider bias.
- In addition, they used different definitions for cancer (i.e. some included only patients with active cancer, while others focused on specific cancer types, etc.), which limits the comparability of their findings, and only a few analysed the effect of cancer across different patient subgroups.

**What this study adds:**

- We conducted a population-based cohort study to analyse the associations between having a prior diagnosis of cancer and the risks of COVID-19 diagnosis, hospitalisation and COVID-19-related deaths from 1 March to 6 May 2020.
- In a population of 4,618,377 adults, we found that cancer was associated with an increased risk of COVID-19 diagnosis (aHR: 1.08; 95% confidence interval [1.05-1.11]); direct COVID-19 hospitalisation (1.33 [1.24-1.43]); and death following a COVID-19 hospitalisation (1.12 [1.01-1.25]).
- These risks were higher for patients recently diagnosed with cancer (within the last year), younger than 70 years, or with haematological cancers. We also found a particularly high risk of COVID-19 hospitalisation and death among patients with lung and bladder cancer.

## Introduction

Cancer is a leading cause of morbidity and death worldwide, with an estimated 19 million new cases and 10 million deaths in 2020.[1] Patients with cancer are often older, have multiple comorbidities, and an impaired immunity due to the cancer itself and cancer therapies, thus increasing their susceptibility to infections.[2] As a result, patients with cancer have been considered a high-risk population for the novel coronavirus disease 2019 (COVID-19) since the beginning of the pandemic.[3] This disease, caused by the severe acute respiratory syndrome coronavirus 2 (SARS-CoV-2), manifests with a varying degree of severity, ranging from asymptomatic to severe disease and death.[4]

Although there is a substantial number of publications addressing the relationship between cancer and COVID-19, these have shown conflicting results.[5] Some studies have found that patients with cancer have an increased risk of COVID-19 infection, hospitalisation and death compared to patients without cancer,[6–9] whereas others have reported null associations.[10–12] The majority of these studies were small, used different criteria to identify patients with cancer (e.g. only active cancers, or solid cancers), and did not include representative samples (i.e. restricted to hospital and/or laboratory-confirmed cases), which limits the generalizability of their findings and increases the risk of selection bias.[13]

Patients with cancer are a highly heterogeneous population that encompasses patients with different features such as cancer type or phases of care since time of diagnosis (e.g., under active treatment, active surveillance or cured). Understanding which patients with cancer are at the highest risk of COVID-19-infection or poor outcomes is essential to inform clinical care and to guide prevention strategies targeting this population. A large, population-based cohort study that includes a heterogeneous cancer population and that captures both COVID-19 incidence and COVID-19-related outcomes could address the limitations of the previous evidence. In this study, we aimed to describe the associations between cancer and the risks of COVID-19 diagnosis, hospitalisation with COVID-19, and COVID-19 related death, overall and by different population subgroups, using real-world data from Catalonia, Spain.

## Methods

### Study design, setting, and data sources

We conducted a population-based cohort study from March 1 2020 until May 6 2020 (last date of data available), using data from the Information System for Research in Primary Care (SIDIAP; www.sidiap.org), a primary care database from Catalonia, a north-eastern region in Spain. Spain has a universal primary care-based health system, in which general practitioners (GPs) are the first point of contact for care. As a consequence, GPs have diagnosed and managed the majority of COVID-19 cases since the beginning of the pandemic.[14] In addition, because GPs are responsible of issuing sick leaves, patients diagnosed with COVID-19 in other settings (e.g., hospital emergency departments) were also bound to contact primary care providers.

The SIDIAP database includes anonymized primary care electronic health records collected since 2006 covering approximately six million people (80% of the population in Catalonia, Spain), and is representative in terms of age, sex, and geographic distribution.[15] SIDIAP includes data on demographics, lifestyle information, and disease diagnoses, among others; and has been linked to SARS-CoV-2 reverse transcription polymerase chain reaction (RT-PCR) test results and hospital records (both from the public sector), as well as to regional mortality data through unique ID linkage. Additionally, SIDIAP has been mapped to the Observational Medical Outcomes Partnership (OMOP) Common Data Model (CDM), allowing us to apply common analytical tools developed by the open-science Observational Health Data Sciences and Informatics (OHDSI) network.[16]

### Study participants

We included all adults (aged 18 years or older) registered in the SIDIAP database as of 1 March 2020 (index date for all participants) with at least one year of prior history observation available. We excluded patients who had a record of a secondary cancer before a record of a primary cancer, patients with a clinical diagnosis or positive test result for COVID-19 prior to index date, and patients hospitalised or living in a nursing home at index date (to include only patients representative of the community population).

### Multistate framework

To address our objectives, we employed a multi-state framework that we have previously utilised to describe the risks of COVID-19 diagnosis, hospitalisation, and death.[17] Multi-state models can be used to describe processes where individuals transition from one health status to another, whilst separating baseline risk and covariate effects associated with each transition.[18] In this study, individuals started the follow-up at the general population and then could transition to three other states: diagnosed with COVID-19 (in an outpatient setting), hospitalised with COVID-19, and death. Six different transitions were possible: from the general population to either diagnosed with COVID-19, hospitalised with COVID-19 (i.e. direct hospitalisation) or death; from diagnosed to either hospitalised with COVID-19 or death; and from hospitalised with COVID-19 to death (Figure 1).

**Figure 1.**
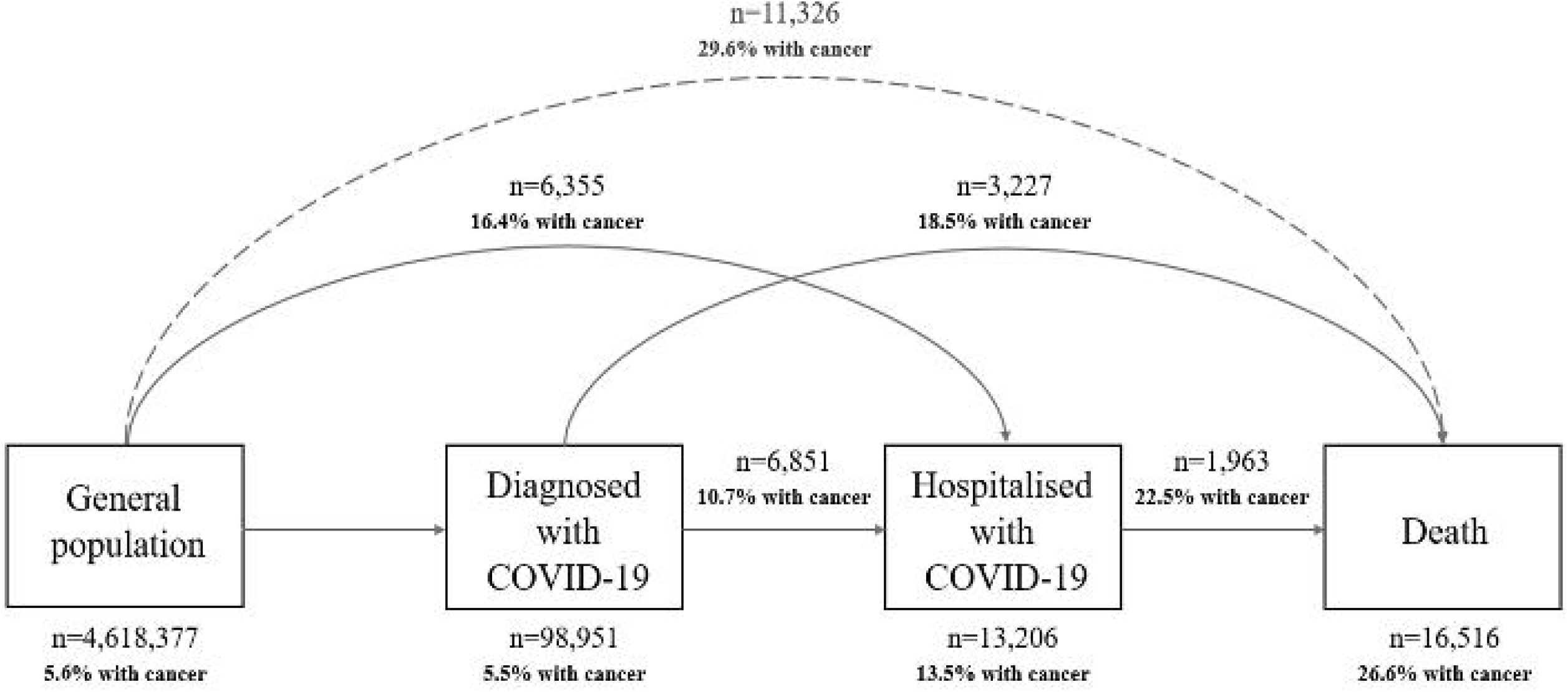
Overview of the multi-state model used in this study.

For all the transitions, individuals were followed until the occurrence of a state of interest, the occurrence of a competing event, or the end of the study period (6 May 2020). Because we were solely interested in COVID-19-related outcomes, we did not model the transition from the general population to death. However, we reported deaths occurring in the general population, which were considered as a competing event.

### Variables

The exposure of interest was cancer, which we defined as any diagnosis of a primary invasive solid or haematological cancer, excluding non-melanoma skin cancer, prior to the index date. We used the International Classification of Diseases, Tenth Revision, Clinical Modification (ICD-10-CM) to identify cancer diagnoses: C00 to C96, except C44 (non-melanoma skin cancer) and C77-C79 (secondary cancers). Cancer types by anatomical location were identified using definitions previously validated in the SIDIAP database.[19] To avoid misclassification of primary cancers, we only considered the earliest cancer type registered for each patient. Patients with cancer were stratified according to the number of years since the diagnosis to the index date into three groups: 1 year, 1-5 years, and ≥5 years.

The covariates of interest were sex, age, smoking status, deprivation, and comorbidities. We extracted participants’ sex and age at index date. Smoking status (never, former, or current smoker) was assigned as the closest assessment to the index date recorded. Deprivation was assessed using the MEDEA deprivation index, which is calculated at the census tract level in urban areas of Catalonia.[20] MEDEA deprivation index is categorised in quintiles, with the first quintile representing the least deprived group and the fifth the most deprived. It also includes a rural category for individuals living in rural areas. Our comorbidities of interest were autoimmune conditions, chronic kidney disease, chronic obstructive pulmonary disease, dementia, heart disease, hyperlipidaemia, hypertension, obesity, and type 2 diabetes. Comorbidities were defined as previously described based on medical diagnosis,[17] and selected due to their relevance to the COVID-19 research field.[21] The definitions for each comorbidity can be consulted in a web application (“Index Event Breakdown” tab) available at https://livedataoxford.shinyapps.io/MultiStateCovidCohorts/.

Our outcomes of interest were an outpatient clinical diagnosis of COVID-19, a hospitalisation with COVID-19, and COVID-19-related death. We defined COVID-19 diagnoses based on a recorded clinical code for COVID-19 disease (ICD-10-CM: B34.2; B97.29). We did not require a positive RT-PCR test result in the definition of COVID-19 diagnoses due to testing restrictions during the first months of the pandemic.[17] We defined hospitalisation with COVID-19 as a hospital admission (with at least one day hospital stay) where the patient had a COVID-19 diagnosis or a positive RT-PCR test result 21 days prior to admission up to three days after admission (to allow for a delay in diagnosis and minimise the risk of including hospital-acquired COVID-19 infections). We extracted deaths (from any cause) from region-wide mortality data, and by doing so we included both deaths during hospitalisation and in the community. Deaths occurring following a COVID-19 event (diagnosis or hospitalisation) were considered as COVID-19-related deaths.

### Statistical analyses

We described participants’ baseline characteristics, participants’ time at risk at each state and numbers of events observed for each transition by cancer status (with or without cancer). To assess the relationship between cancer and the risk of transitioning to a subsequent state in the multistate model, we estimated adjusted cause-specific hazard ratios (aHRs), with 95% confidence intervals (CIs), using Cox proportional hazard regressions for each transition.

First, we estimated models for all patients with cancer compared to patients without cancer adjusting for age, sex, the MEDEA deprivation index, smoking status, and all the comorbidities of interest (main models). We used a directed acyclic graph to guide decisions on the control for confounding (Figure S1).[22] To check the proportional hazard assumptions for the variables included in the models, we visually inspected log-log survival curves. Missing data were handled as an additional category. Non-linearity in age and risks of transition was considered by fitting models with age as a linear term, with a polynomial of degree 2 (i.e. quadratic), and with restricted cubic splines (with 3, 4, or 5 knots).[23] We1 calculated the Bayesian Information Criterion (BIC) for each of those models and we selected the models with the lowest BIC values.

Second, we estimated the relationship between cancer and COVID-19 outcomes adjusting for age and sex; and adjusting for age, sex, the MEDEA deprivation index, and smoking status. Third, we further estimated our main models separately for <1-year, 1-5-years, and >5-years cancer patients, and stratified these models by sex (women or men), age (<70 and ≥70 years, 70 years was the median age of patients with cancer), cancer type (haematological or solid cancer, as well as by solid cancer types). All models were relative to patients without cancer (cancer-free).

As sensitivity analyses, we re-estimated our main models: 1) stratifying by calendar time for transitions in which the proportionality assumption was violated; 2) restricting participants to never-smokers, to avoid residual confounding by smoking; and 3) after performing a multiple imputation of missing data (smoking status and MEDEA deprivation index) using predictive mean matching, with 5 imputations drawn. We also compared baseline characteristics of patients with and without missing data using standardized mean differences (SMD). We considered SMD ≥|0.1| as a meaningful difference in the distribution of a given characteristic between the two groups.[24]

We used R version 3.6 for data analysis and visualization. The R packages used in the analysis included mstate,[25] and rms.[26] The analytic code is available at https://github.com/SIDIAP/COVID-19-cancer-multi-state. This study was approved by the Clinical Research Ethics Committee of the IDIAPJGol (project code: 20/070-PCV).

### Public and patient involvement statement

Participants of this study were not involved in setting the research question or the outcome measures, nor were they involved in the design or implementation of the study. No patients were asked to advise on interpretation or writing of results

## Results

### Population included

A total of 4,618,377 adults were included. We excluded 104,022 individuals with less than a year of prior observation history; 1,496 with a record of a secondary cancer before a record of a primary cancer; 303 with a COVID-19 diagnosis or positive SARS-CoV-2 test before index date; 40,421 living in a nursing home, and 1,138 hospitalised at the index date (Figure S2). Baseline characteristics of the population included are summarised in Table 1. In total, 260,667 (5.6%) patients had a prior diagnosis of cancer. Of these, 167,053 (64.1% of the cancer population) were diagnosed ≥5 years; 72,033 (27.6%) 1-5 years; and 21,581 (8.3%) <1 year prior to the index date. Compared to cancer-free patients, those with cancer were older, more frequently former smokers and living in the least deprived areas of Catalonia. In addition, they had a higher burden of comorbidities, especially cardiovascular conditions (e.g., 27.4% had heart disease vs. 10.2% in cancer-free patients). When stratifying patients by age categories, we observed that the burden of comorbidities increased with age for both groups (Figure S3). Among patients with cancer, 239,030 (91.7%) and 21,637 (8.3%) had a solid and haematological cancer, respectively. The most frequent solid cancer types were breast (n=58,611, 22.5%), prostate (37,141, 14.2%), colorectal (36,071, 13.8%) and bladder (20,592, 7.9%).

**Table 1.**
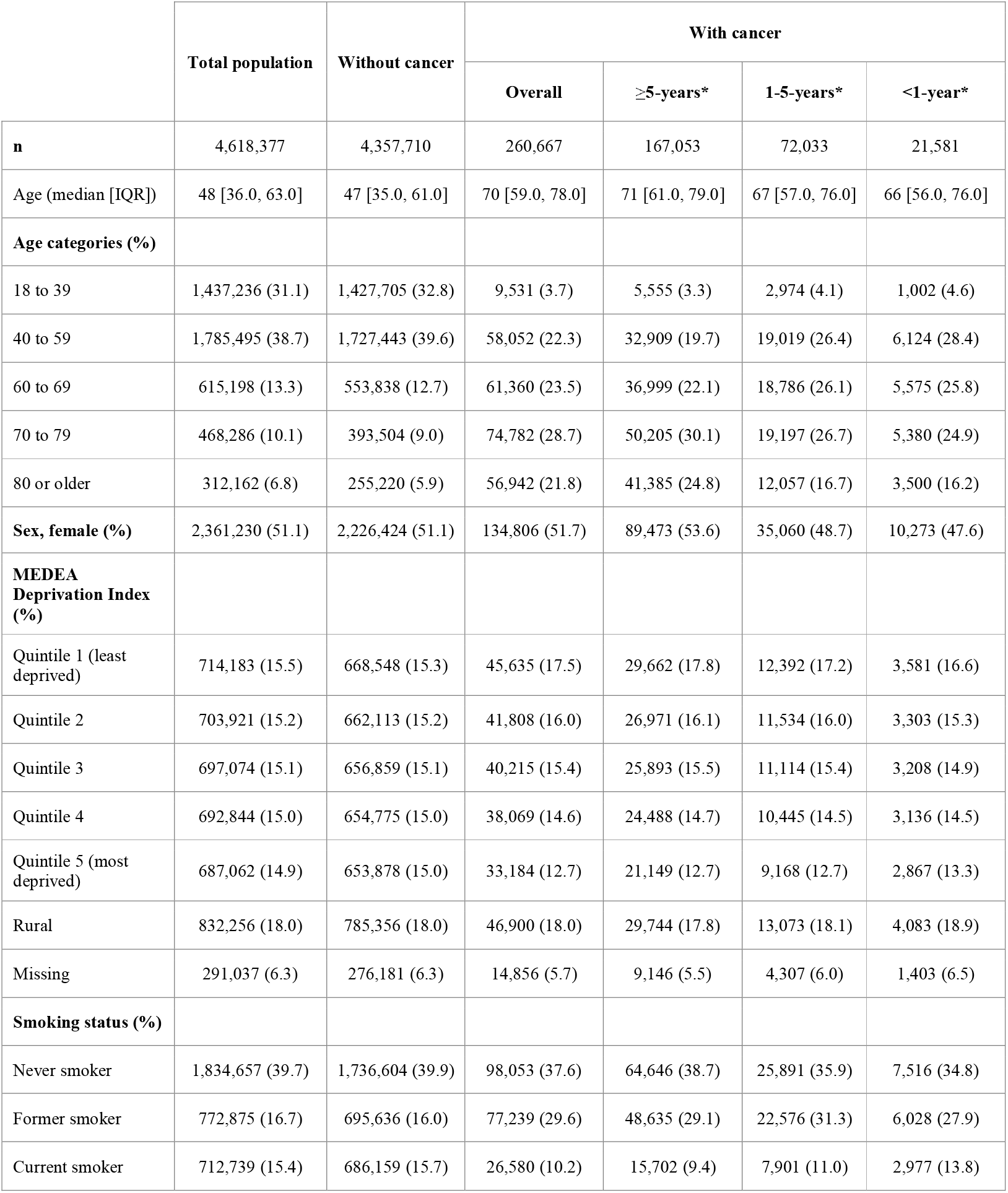
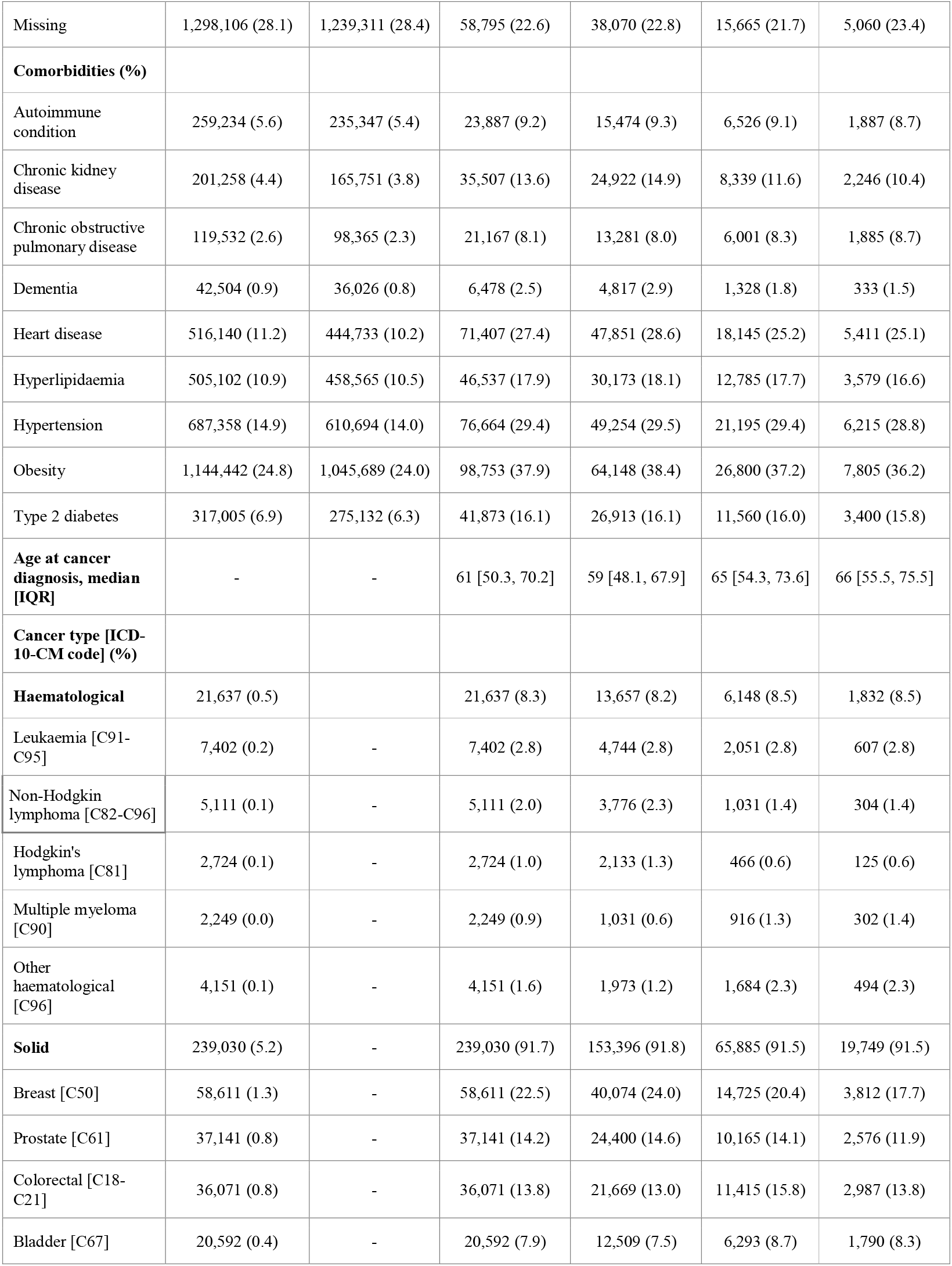
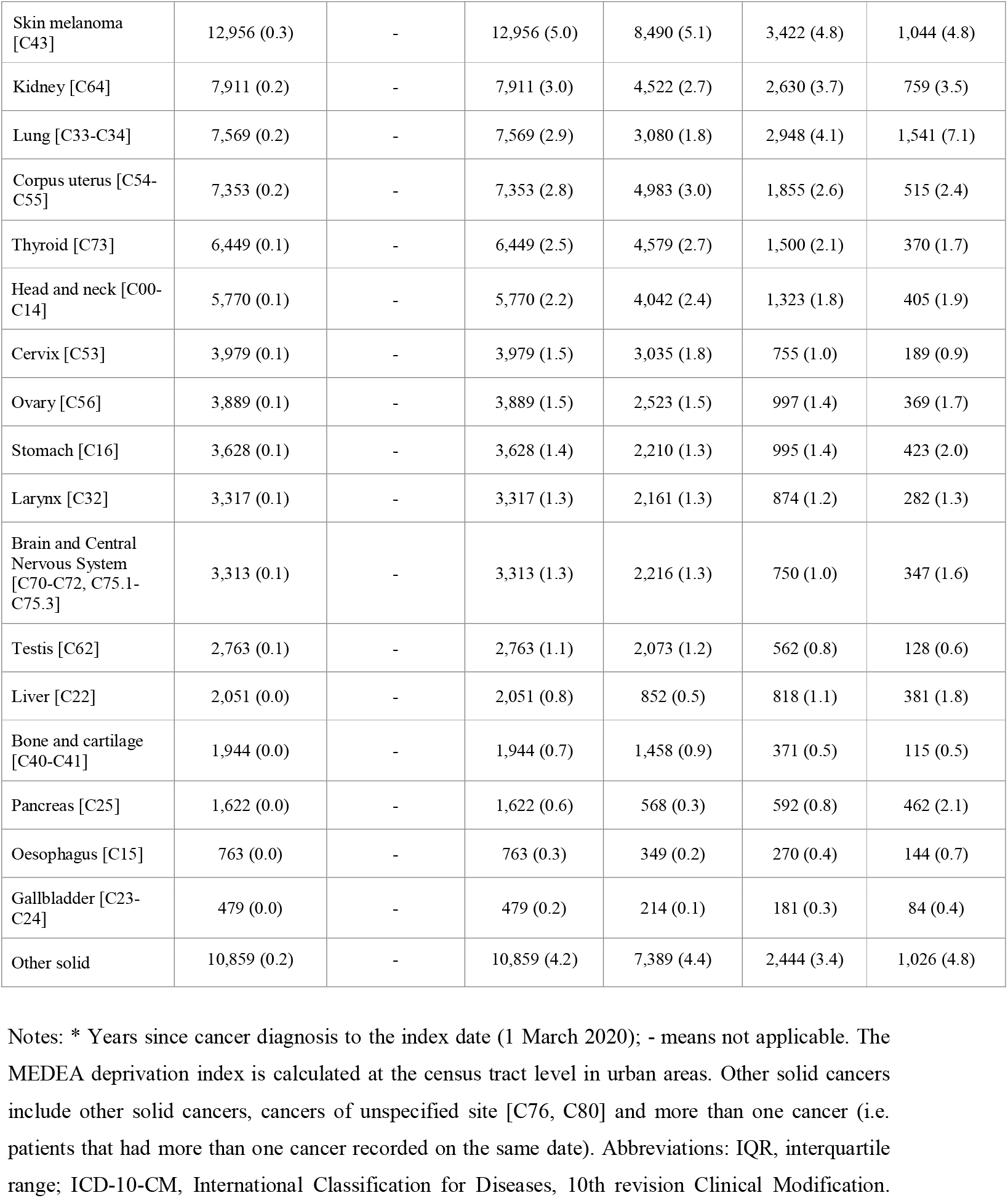
Baseline characteristics of the population included, by cancer status.

### Occurrence of COVID-19 outcomes

Among the general population, 98,951 (2.1% cumulative incidence (CI) at 67 days) individuals were diagnosed with COVID-19; 6,355 (0.1% CI) were directly hospitalised with COVID-19 and 11,326 (0.25% CI) died without a COVID-19 diagnosis/hospitalisation (Figure 1, Table 2). Among individuals diagnosed with COVID-19, 6,851 (7.2% CI at 45 days) were hospitalised and 3,227 (3.9% CI) died without a hospitalisation. Among those hospitalised, 1,963 (18% CI at 45 days) died.

**Table 2.**
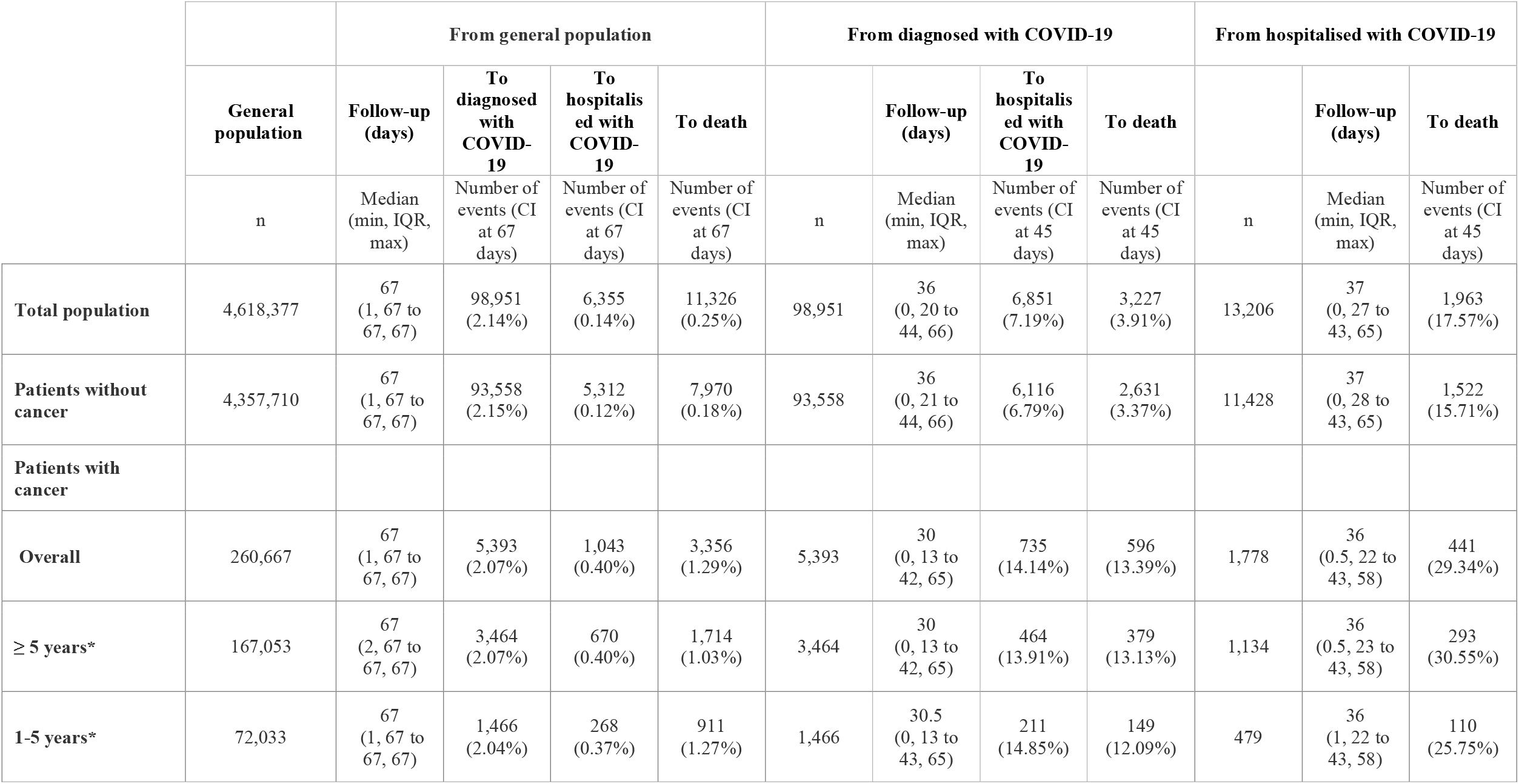
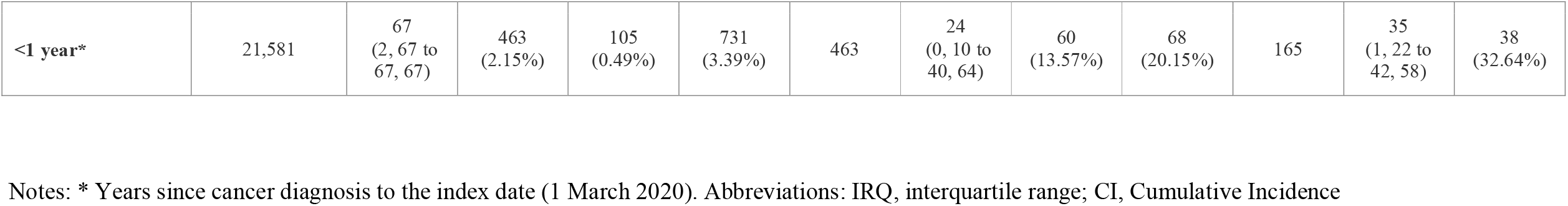
Time at risk, absolute number of events, and cumulative incidence, by cancer status.

Among the total cancer population (n=260,667), 5,393 (2.1% CI at 67 days) patients were diagnosed with COVID-19; 1,043 (0.4%) were directly hospitalised with COVID-19 and 3,356 (1.3%) died without a COVID-19 diagnosis/hospitalisation. Among those diagnosed with COVID-19, 735 (14.1% CI at 45 days) were subsequently hospitalised and 596 (13.4%) died without a hospitalisation. Among those hospitalised, 441 (29.3% CI at 45 days) died. Descriptive characteristics by state and transition are shown in Table S1. Briefly, individuals diagnosed/hospitalised with COVID-19, as well as having a COVID-19-related death, were older, more frequently male and former smokers, and had more comorbidities than the general population.

### Risks of COVID-19 diagnosis, hospitalisation and death among patients with cancer

Compared to cancer-free patients, those with cancer had an increased risk of COVID-19 diagnosis (overall aHR: 1.08; 95% CI [1.05-1.11]); direct COVID-19 hospitalisation (1.33 [1.24-1.43]); and death following a COVID-19 hospitalisation (1.12 [1.01-1.25]) (Figure 2). Models using different adjustment strategies showed similar results to our main models (Figure S4).

**Figure 2.**
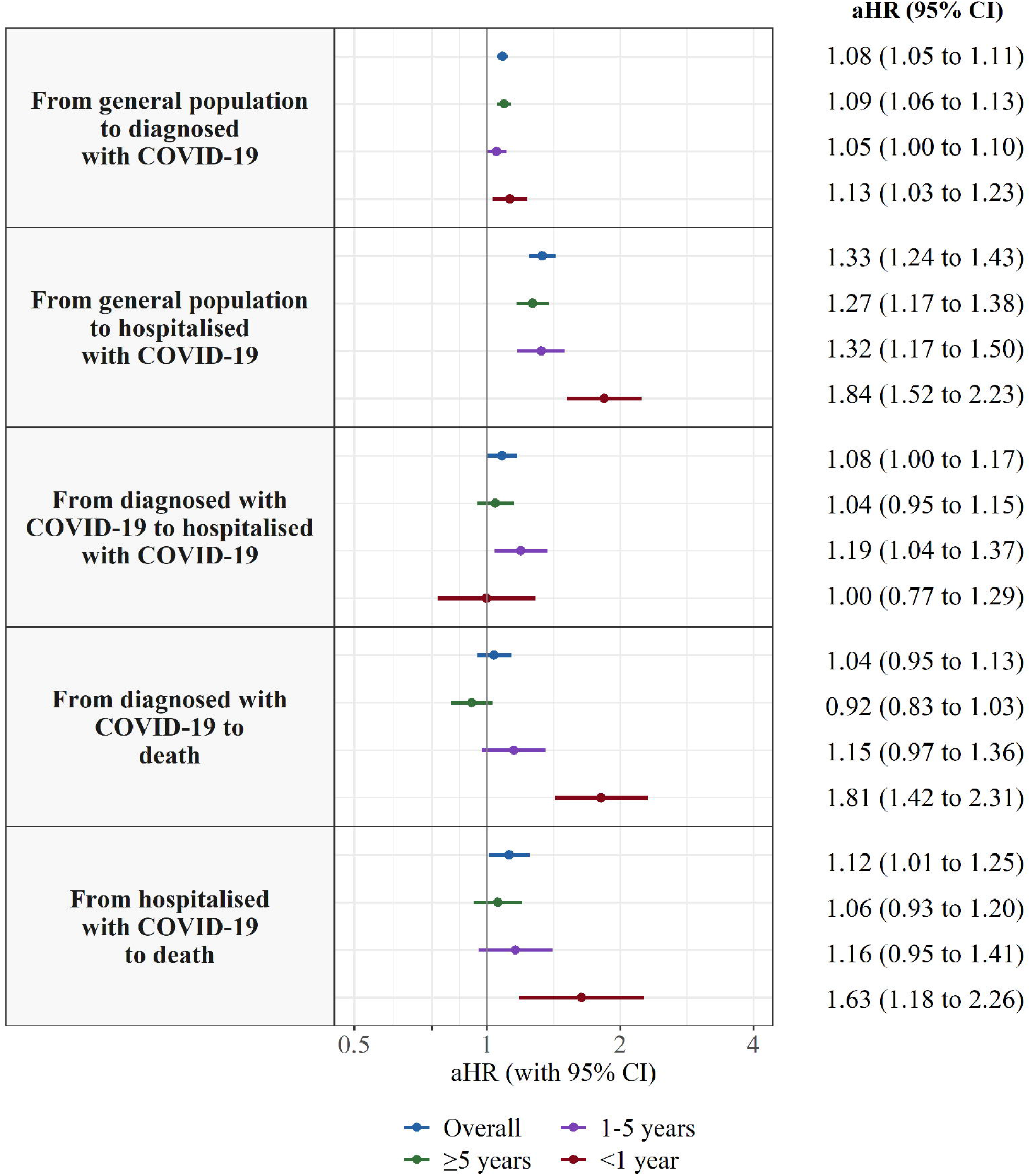
Adjusted Hazard Ratios of COVID-19 outcomes in patients with cancer compared to patients without cancer, overall and by years since cancer diagnosis. Notes: Models are adjusted for age, sex, the MEDEA deprivation index, smoking status, and comorbidities (autoimmune conditions, chronic kidney disease, chronic obstructive pulmonary disease, dementia, heart disease, hyperlipidaemia, hypertension, type 2 diabetes, and obesity). Abbreviations: aHR, adjusted Hazard Ratio; CI, Confidence Interval.

In models stratified by years since cancer diagnosis, the risk of COVID-19 diagnosis was similar in <1-year, 1-5-year and ≥5-year cancer patients (Figure 2). As for the risk of direct COVID-19 hospitalisation, <1-year cancer patients had the highest risk (1.84 [1.52-2.23]), followed by 1-5-year cancer patients (1.32 [1.17-1.50]) and ≥5-year cancer patients (1.27 [1.17-1.38]). Increased risk of COVID-19-related death remained significant only in <1-year cancer patients, for both deaths following a COVID-19 diagnosis (1.81[1.42-2.31]) and following a COVID-19 hospitalisation (1.63 [1.18-2.26]).

Overall, in models stratified by sex, the associations between cancer and risk of COVID-19 diagnosis and death (following a diagnosis/hospitalisation) were moderately stronger in men, whereas the associations with risk of direct hospitalisation were moderately stronger in women (Figure 3, Table S2). In models stratified by age, we found a stronger association between cancer and COVID-19 outcomes in the subgroup of patients aged <70 years compared to those aged ≥70 years, aside from the risk of COVID-19 diagnosis (Figure 3, Table S3). Age differences were more pronounced in <1-year cancer patients. Additionally, the associations between cancer and COVID-19-related death (either following a COVID-19 diagnosis or a hospitalisation) were only significant in the subgroup of patients aged <70 years. For example, the overall aHR for death following hospitalisation was 1.49 [1.10-2.01] in <70-years patients and 1.07 [0.95-1.20] in ≥70-years patients. In <1-year cancer patients, the aHR was 4.58 [2.47-8.50] in <70-years patients and 1.30 [0.88-1.90] in ≥70-years patients.

**Figure 3.**
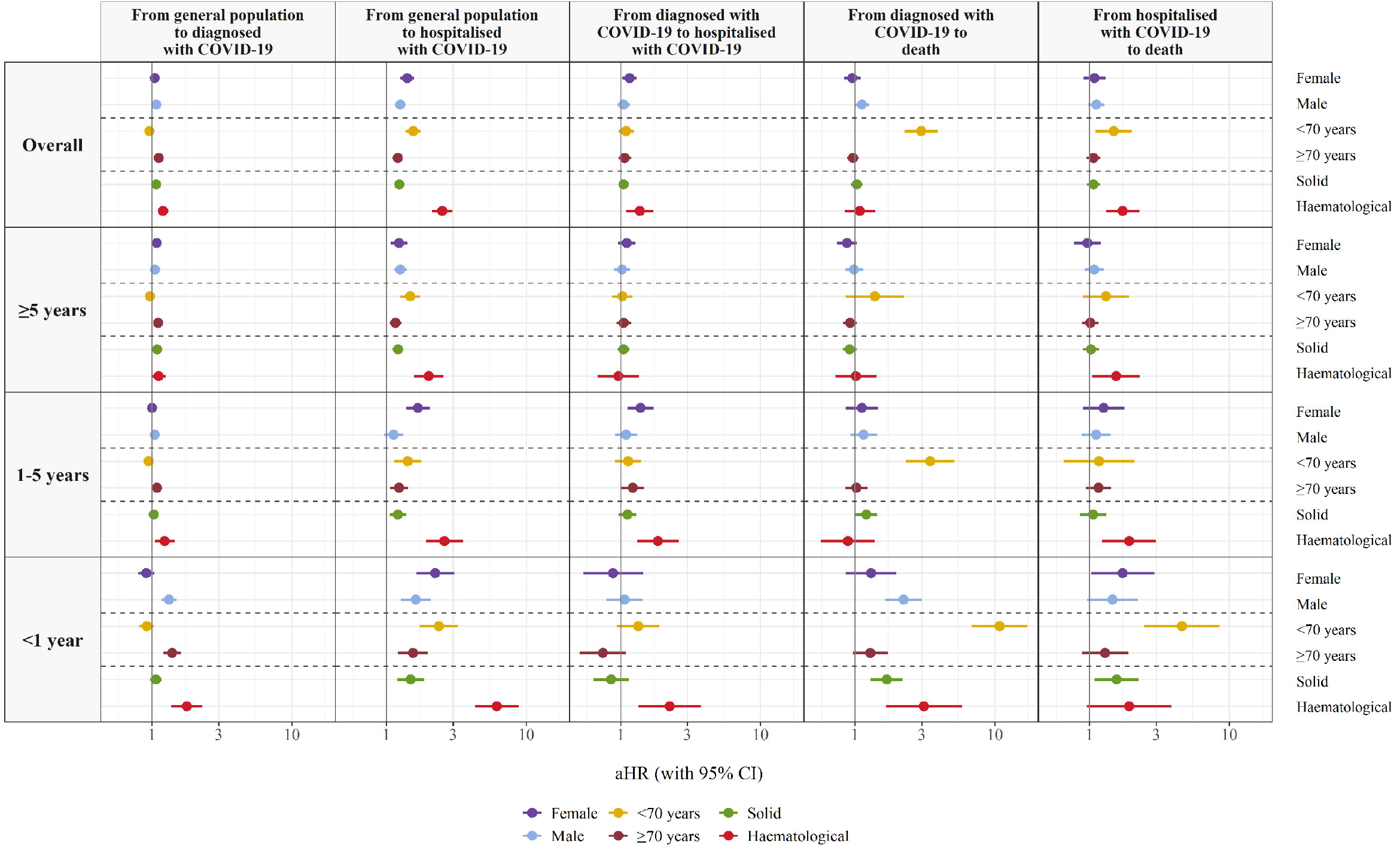
Adjusted Hazard Ratios of COVID-19 outcomes in patients with cancer (overall and by years since cancer diagnosis) compared to patients without cancer, stratified by sex, age, and cancer type (solid or haematological) Notes: Models are adjusted for sex (excepting models stratified by sex), age, the MEDEA deprivation Index, smoking status, and comorbidities (autoimmune conditions, chronic kidney disease, chronic obstructive pulmonary disease, dementia, heart disease, hyperlipidaemia, hypertension, type 2 diabetes, and obesity). Abbreviations: aHR, adjusted Hazard Ratio; CI, Confidence Interval

When stratifying patients by haematological or solid cancers, those with haematological cancers had a higher risk of COVID-19 outcomes (Figure 3, Table S4). These differences were more pronounced in <1-year cancer patients. For example, the overall aHR for having a direct COVID-19 hospitalisation was 2.51 [2.12-2.98] for patients with haematological cancers and 1.24 [1.15-1.33] for those with solid cancers. Among <1-year cancer patients, aHR were 6.18 [4.31-8.86] for haematological cancers and 1.49 [1.19-1.87] for solid cancers. Patients with haematological cancers also had an increased risk of COVID-19 hospitalisation following an outpatient diagnosis (overall 1.37 [1.10-1.71]; <1-year cancer patients: 2.24 [1.34-3.76]).

We also estimated the associations between cancer and COVID-19 outcomes by solid cancers. (Figure S5, Table S5). Due to small samples, models were estimated for breast, prostate, colorectal, bladder, and lung cancer; overall and for <5-years (<1-year and 1-5-year categories combined) and ≥5-years cancer patients. Four cancer types were associated with having a direct COVID-19 hospitalisation: breast (1.30 [1.10-1.54]), colorectal (1.28 [1.10-1.49]), bladder (1.50 [1.26-1.79]) and lung (1.53 [1.13-2.08]) cancer; these associations were stronger in <5-year cancer patients. Lung cancer was associated with death following a COVID-19 diagnosis (1.68 [1.06-2.64]), with a stronger association in <5-year cancer patients (2.57 [1.49-4.46]). Bladder cancer was associated with death following a COVID-19 hospitalisation only in <5-year cancer patients (1.70 [1.11-2.60]).

### Sensitivity analysis

The assumption of proportionality was violated for age and years since cancer diagnosis for the risk of COVID-19 diagnosis (Figure S6). Thus, we stratified our model by years since cancer diagnosis and calendar time (Figure S7). The overall association was similar in March and April. However, in <1-year cancer patients, cancer was associated with a significant increased risk of COVID-19 diagnosis in April (1.41 [1.23-1.60]) but not in March (0.91 [0.80-1.05]).

In models restricted to never smokers (n=1,834,657), the results were similar to those including all the population (Figure S8). Patients with missing data (n=1,502,442) were younger and had fewer comorbidities than patients without missing data, but the distribution of cancer types was similar in both groups (Table S6). Despite these differences, imputed models showed similar results to the main models (Figure S9).

## Discussion

In this population-based cohort study including 4,618,377 adults, a prior diagnosis of cancer was associated with an increased risk of COVID-19 outpatient (clinical) diagnosis, direct COVID-19 hospitalisation (without a prior outpatient diagnosis), and COVID-19-related death during the first wave of the COVID-19 pandemic in Catalonia, Spain. Overall, these associations were stronger in patients with a recent cancer diagnosis (<1 year), younger than 70 years, and with haematological cancers. Lung and bladder cancers were also associated with higher risk of COVID-19 hospitalisation and death.

This study has several strengths. First, we used prospective data from a large and representative population covering almost all the population in Catalonia and we included a heterogeneous cancer population. Secondly, by including patients with a clinical COVID-19 diagnosis, we avoided selection bias due to testing restrictions, or to (hypothetically) different testing patterns (i.e., higher rates of testing in patients with cancer), although some cases might be false positives. Thirdly, we performed our analysis across different cancer population groups, allowing us to identify those at highest risk of poor COVID-19 outcomes. Finally, our results were robust after restricting participants to never smokers and after multiple imputation of missing data, which lends credibility to our findings.

However, this study also has weaknesses. First, we did not have information on cancer stage nor specific-cancer therapy receipt, and used instead years since cancer diagnosis as a proxy for active/inactive cancer. We also did not have information on the cause of death and considered as COVID-19-related deaths those occurring following a COVID-19 state. However, in patients with cancer, occurrence of death was substantially higher in those diagnosed (11.1%) and hospitalised (24.8%) with COVID-19 than in those without COVID-19 (1.3%), which suggests that we did capture deaths due to COVID-19. In addition, the proportion of deaths among hospitalised patients was in line with prior studies.[27] On the other hand, we cannot discard that some deaths in the general population might have occurred in undiagnosed COVID-19 cases, especially at the beginning of the pandemic. Secondly, due to the nature of our database, our results are not representative of asymptomatic or pauci-symptomatic COVID-19 cases that did not seek medical care. Finally, routinely collected data often raises concerns about data quality and some conditions, including cancer itself, may have been incompletely or inaccurately recorded. However, we used previously validated cancer codes,[19] and we included only individuals with at least one year of prior history available to comprehensively capture baseline characteristics.

Prior studies investigating the risk of contracting SARS-CoV-2 in patients with cancer have reported conflicting results.[6,10,28,29] Even though we did not analyse the risk of COVID-19 infection per se, patients with cancer had a modestly increased risk of having an outpatient COVID-19 diagnosis, which was higher in <1-year cancer patients with haematological cancers. This is consistent with two studies from the United States (US) showing an increased risk of infection in patients with cancer, which was higher in those recently diagnosed and/or with haematological cancers.[6,28] Increased risk of diagnosis could be related to higher levels of interaction with healthcare services among patients with cancers (thus, higher risk of being diagnosed but also higher exposure to healthcare-associated infections), and to factors related to the cancer itself and/or cancer therapies (e.g. haematological cancers, as well as treatment-related immunosuppression, thus increasing risk of infection).[30]

Patients with cancer have also been reported to be at increased risk of COVID-19 severity, including hospitalisation and death.[6–9] We found that cancer was associated with a higher risk of direct hospitalisation with COVID-19, regardless of years since cancer diagnosis; whereas only 1-5-year cancer patients had a higher risk of subsequent hospitalisation (following an outpatient diagnosis). This counterintuitive finding could be explained by differences in care-seeking behaviours and/or in the clinical presentation of COVID-19. On the one hand, patients with cancer (especially those recently diagnosed) could be more prone to seek care directly at the hospital level than the general population.[31] On the other hand, these patients might have a higher risk of rapidly developing severe COVID-19 symptoms due to their impaired immunity. It is worth noting that although <1-year cancer patients had the highest risk of hospitalisation, this association remained significant in >5-year cancer patients (which mostly represent cancer survivors). This is consistent with a study showing that cancer survivors have higher risks of hospitalisation and death from influenza than cancer-free patients,[32] and could be related to long-term effects on the immune system of cancer therapies.

Conversely, the risk of COVID-19-related death was only significantly higher in <1-year cancer patients. This could be related to cancer treatments; however, while some studies have shown that active cancer therapies increase the risk of COVID-19 death,[9] others have not.[8,33] These studies included different populations, cancer types, or considered all different cancer therapies combined, which might have a different impact on COVID-19 outcomes. For instance, two meta-analyses reported an association between recent chemotherapy and increased COVID-19-related death, but a null association with recent surgery, radiotherapy, immunotherapy and targeted therapies.[34,35]

We found that the associations between cancer and direct hospitalisation and COVID-19-related deaths were more pronounced in patients younger than 70 years or with haematological cancers. Given that age is strongly associated with severe COVID-19 outcomes, cancer in older patients might not have a significantly worse impact as compared to cancer-free patients. In a study including 1,187 patients with solid cancers and COVID-19, younger patients (<60 years) were also those with the highest risk of in-hospital mortality when compared to cancer-free patients.[36] Furthermore, increasing evidence shows that patients with haematological cancers have a higher risk of poor COVID-19 outcomes.[6,7,9] The OpenSAFELY study reported an association between cancer and increased COVID-19 death, which was stronger in <1-year cancer patients and in those with haematological cancers.[7] Estimated aHR for <1-year cancer patients were similar to ours for death following a COVID-19 diagnosis, with an aHR of 1.72 [1.50-1.96] (vs 1.69 [1.30-2.19] in our study) for solid cancer patients; and an aHR of 2.80 [2.08-3.78] (vs 3.11 [1.67-5.81]) for haematological cancer patients. We also found a higher risk of hospitalisation and COVID-19-related death for lung and bladder cancers, both of which are strongly linked to tobacco smoking. While lung cancer has already been associated with poor COVID-19 outcomes,[37] to our knowledge this study is the first showing an association with bladder cancer. However, these findings should be interpreted with caution considering the small sample sizes, which prevented us from performing analysis restricted to never smokers by specific cancer types.

This population-based cohort study including a heterogeneous cancer population provides a comprehensive analysis of the associations between cancer and COVID-19 outcomes during the first wave of the pandemic in a Southern European region. Cancer was associated with an increased risk of COVID-19 diagnosis, hospitalisation, and COVID-19-related death, with higher risks for patients diagnosed with cancer within the year prior, as well as those younger than 70 years and those with haematological cancers. Research is needed to address potential risk differences by specific cancer types, such as lung or bladder cancer, as well as to analyse the effect of subsequent COVID-19 waves. Notwithstanding that, our results highlight that patients with cancer are a vulnerable population for COVID-19. These patients, as well as their caregivers, should be prioritised in preventive strategies, including vaccination campaigns and continued non-pharmaceutical interventions.

## Data Availability

In accordance with current European and national law, the data used in this study is only available for the researchers participating in this study. Thus, we are not allowed to distribute or make publicly available the data to other parties. However, researchers from public institutions can request data from SIDIAP if they comply with certain requirements. Further information is available online (https://www.sidiap.org/index.php/menu-solicitudesen/application-proccedure) or by contacting Anna Moleras.

## Acknowledgements

We would like to acknowledge the patients who suffered from or died of this devastating disease, and their families and carers. We would also like to thank the healthcare professionals involved in the management of COVID-19 during these challenging times, from primary care to intensive care units in the Catalan healthcare system.

Voldríem reconèixer i tindre un record per tots els pacients que han patit i els que han mort per la COVID-19. Volem també agrair tots els professionals sanitaris que han diagnosticat i tractat aquesta malaltia al sistema català de salut, des dels centres d’atenció primària fins a les unitats de cures intensives.

Queremos reconocer y recordar a todos los pacientes que han sufrido y fallecido por la COVID-19. Queremos también agradecer a todos los profesionales sanitarios que han diagnosticado y tratado esta enfermedad en el sistema catalán de salud y en España, desde los centros de salud hasta las unidades de cuidados intensivos.

The analysis has made use of a range of open-source, free available tools provided by the OHDSI community. The phenotypes used in this study were developed or adapted from work performed during the OHDSI COVID-19 studyathon. The mapping of data to the OMOP CDM has been supported by a taskforce from EHDEN.

## Ethical approval

This study was approved by the Clinical Research Ethics Committee of the IDIAPJGol (project code: 20/070-PCV)

## Transparency statement

ER and TDS as guarantors of the manuscript affirm that the manuscript is an honest, accurate, and transparent account of the study being reported; that no important aspects of the study have been omitted; and that any discrepancies from the study as planned have been explained.

## Author Contributions

SFB, MA, and TDS mapped source data to the OMOP CDM. ER, EB, AP, and TDS led the data analysis. ER performed the literature review. ER wrote the first draft with insightful contributions from MR and TDS. All authors were involved in the study conception and design, interpretation of the results, manuscript preparation, and approved the final version of the manuscript. The corresponding author attests that all listed authors meet authorship criteria and that no others meeting the criteria have been omitted.

## Disclaimer

Where authors are identified as personnel of the International Agency for Research on Cancer and World Health Organization, the authors alone are responsible for the views expressed in this Article and they do not necessarily represent the decisions, policy, or views of the International Agency for Research on Cancer and World Health Organization.

