## Supplementary for "Cancer and the risk of COVID-19 diagnosis, hospitalisation, and death: a population-based multi-state cohort study including 4,618,377 adults in Catalonia, Spain"

Elena Roel, doctoral student, ^1,2^ Andrea Pistillo, statistician,^1^ Martina Recalde, doctoral student,^1,2^ Sergio Fernández-Bertolín, data scientist,^1^ María Aragón, SIDIAP coordinator,^1^ Isabelle Soerjomataram, scientist,^3^ Mazda Jenab, scientist,^3^ Diana Puente, senior epidemiologist,^1^ Daniel Prieto-Alhambra, Professor of pharmaco- and device epidemiology,^4^ Edward Burn, postdoctoral researcher,^1,4^ Talita Duarte-Salles, senior epidemiologist.^1^*

1. Fundació Institut Universitari per a la recerca a l’Atenció Primària de Salut Jordi Gol i Gurina (IDIAPJGol), Gran Via Corts Catalanes, 587 àtic, 08007, Barcelona, Spain

2. Universitat Autònoma de Barcelona, 08193, Bellaterra (Cerdanyola del Vallès), Barcelona, Spain

3. International Agency for Research on Cancer (IARC-WHO), 150 Cours Albert Thomas, 69008 Lyon, France

4. Centre for Statistics in Medicine, Nuffield Department of Orthopaedics, Rheumatology, and Musculoskeletal Sciences, University of Oxford, Botnar Research Centre, Windmill Road, Oxford, OX3 7LD, UK.

*Corresponding author

Talita Duarte-Salles

Fundació Institut Universitari per a la recerca a l'Atenció Primària de Salut Jordi Gol i Gurina (IDIAPJGol)

Gran Via Corts Catalanes, 587 àtic

08007 Barcelona - Spain

**Supplementary**

Figure S1. Directed Acyclic Graph used to adjust the Cox proportional hazard models to assess the relationship between cancer and COVID-19 outcomes	2

Figure S2. Flowchart with the inclusion and exclusion criteria of the study population	3

Figure S3. Prevalence of comorbidities among the general population, by age and cancer status	4

Table S1. Descriptive characteristics of the population included, by state and transition	5

Figure S4. Hazard Ratios of COVID-19 outcomes in patients with cancer compared to patients without cancer, using different adjustment strategies.	9

Table S2. Adjusted Hazard Ratios of COVID-19 outcomes in patients with cancer (overall and by years since cancer diagnosis) compared to patients without cancer, stratified by sex.	10

Table S3. Adjusted Hazard Ratios of COVID-19 outcomes in patients with cancer (overall and by years since cancer diagnosis) compared to patients without cancer, stratified by age.	11

Table S4. Adjusted Hazard Ratios of COVID-19 outcomes in patients with cancer (overall and by years since cancer diagnosis) compared to patients without cancer, stratified by solid or haematological cancer.	12

Figure S5. Adjusted Hazard Ratios of COVID-19 outcomes in patients with cancer (overall and by years since the cancer diagnosis) compared to patients without cancer, stratified by solid cancer type.	13

Table S5. Adjusted Hazard Ratios of COVID-19 outcomes in patients with cancer (overall and by years since cancer diagnosis) compared to patients without cancer, stratified by solid cancer type.	15

Figure S6. Log-log plots for risk of having an outpatient COVID-19 diagnosis.	17

Figure S7. Adjusted Hazard Ratios for having an outpatient COVID-19 diagnosis in patients with cancer (overall and by years since the cancer diagnosis) compared to patients without cancer, stratified by month	19

Figure S8. Adjusted Hazard Ratios of COVID-19 outcomes in patients with cancer compared to patients without cancer, by population included (all and restricting participants to never smokers)	20

Table S6. Comparison of baseline characteristics among individuals with complete information on all covariates vs individuals with missing information on the MEDEA deprivation index and/or smoking status.	21

Figure S9. Adjusted Hazard Ratios of COVID-19 outcomes in patients with cancer compared to patients without cancer, after multiple imputation of missing data on the MEDEA deprivation index and/or smoking status	**24**

### Figure S1. Directed Acyclic Graph used to adjust the Cox proportional hazard models to assess the relationship between cancer and COVID-19 outcomes

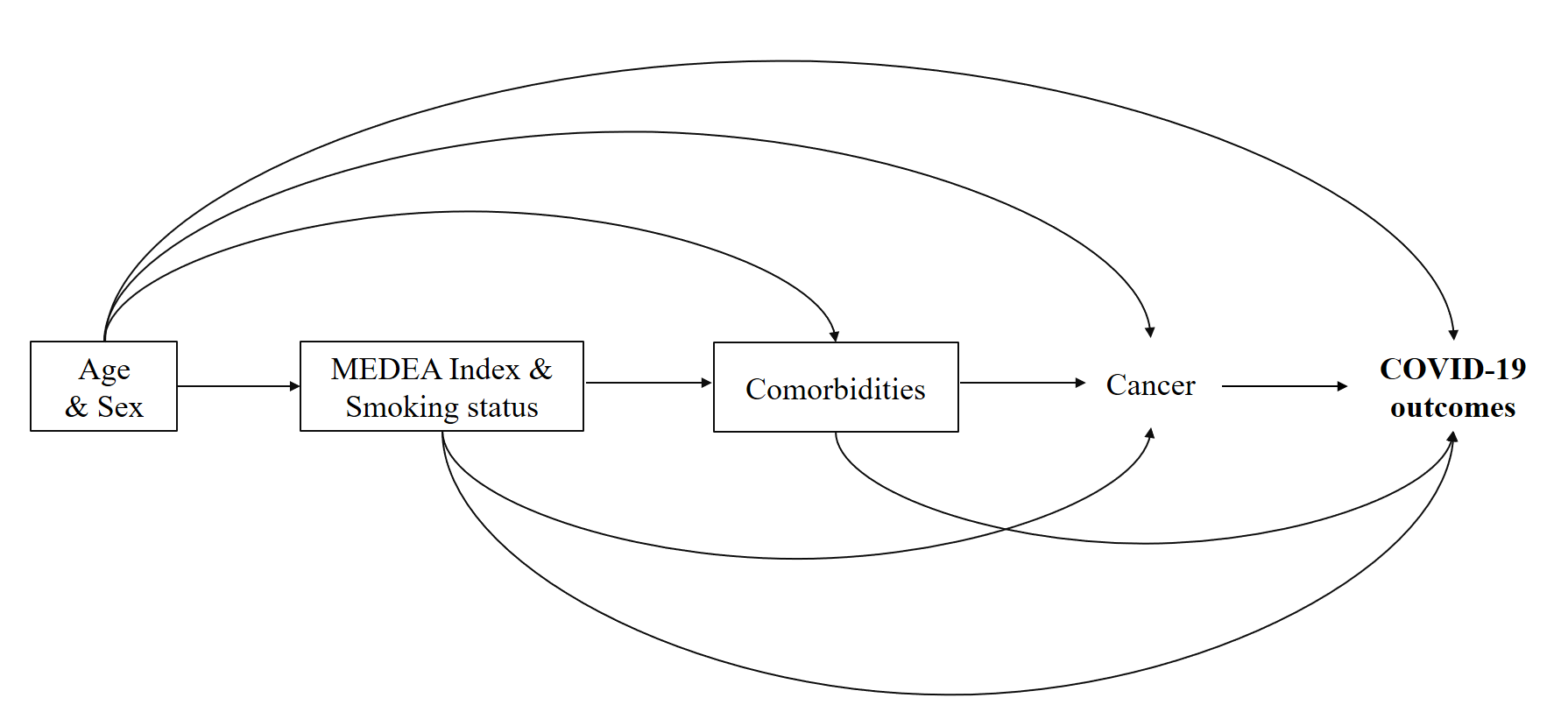

Notes: MEDEA is an index of deprivation calculated at the census tract level in urban areas. The comorbidities considered are autoimmune conditions, chronic kidney disease, chronic obstructive pulmonary disease, dementia, heart disease, hyperlipidaemia, hypertension, obesity, and type 2 diabetes.

### Figure S2. Flowchart with the inclusion and exclusion criteria of the study population

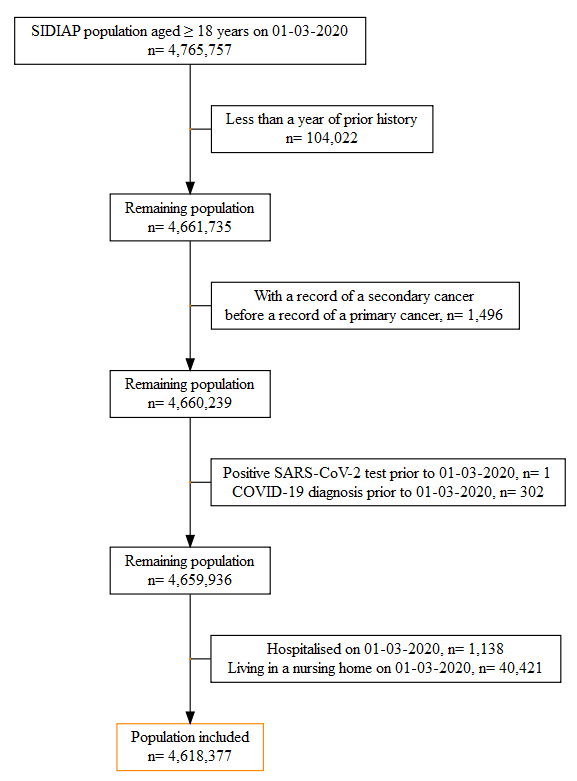

Abbreviations: SIDIAP: Information System for Research in Primary Care; SARS-CoV-2: Severe Acute Respiratory Syndrome Coronavirus 2

### Figure S3. Prevalence of comorbidities among the general population, by age and cancer status

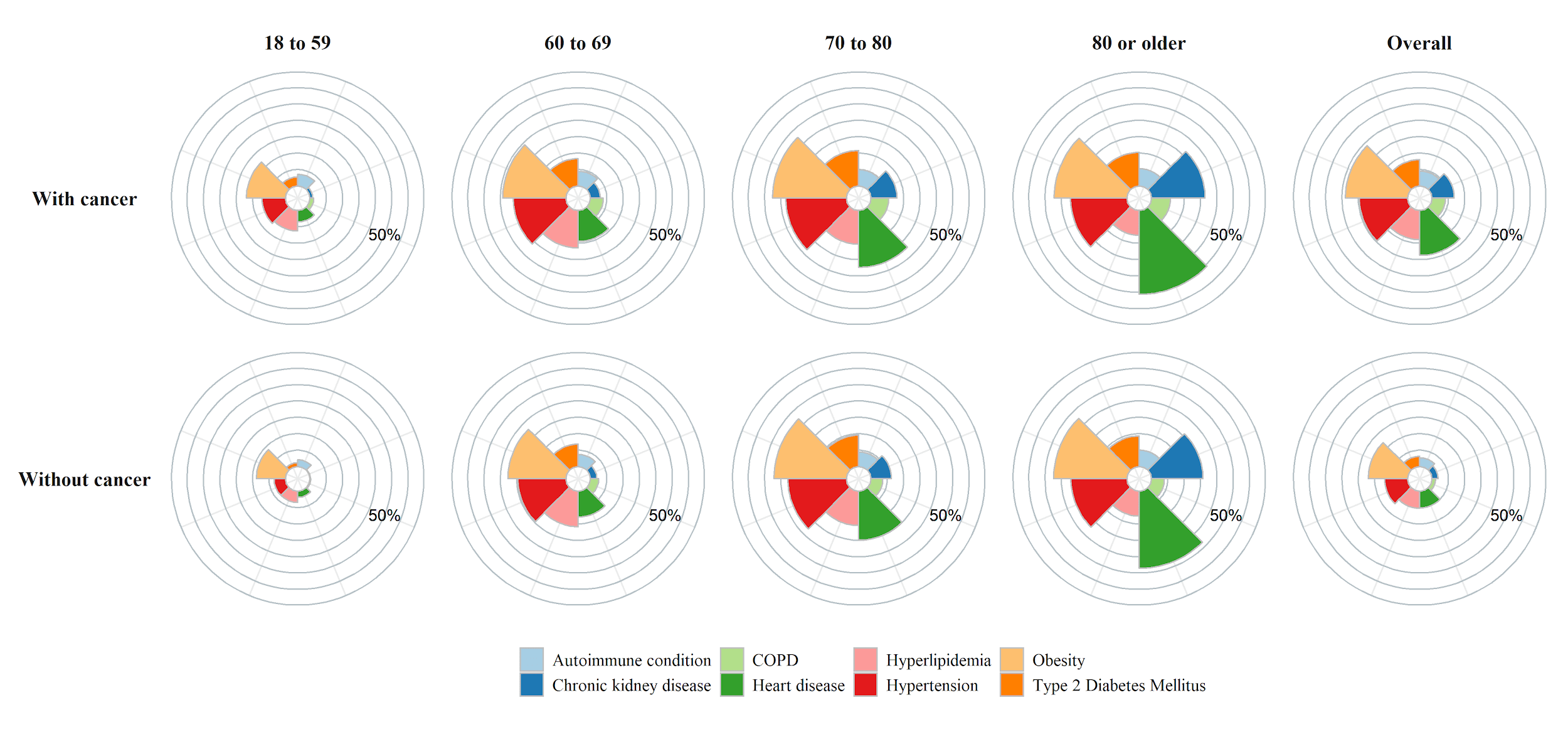

Abbreviations: COPD, Chronic Obstructive Pulmonary Disease

### Table S1. Descriptive characteristics of the population included, by state and transition

|  |  | **Among general population** | | | **Among diagnosed with COVID-19** | | **Among hospitalised with COVID-19** |
| --- | --- | --- | --- | --- | --- | --- | --- |
|  | **General population** | **Diagnosed with COVID-19** | **Hospitalised with COVID-19** | **Death** | **Hospitalised with COVID-19** | **Death** | **Death** |
| n | 4,618,377 | 98,951 | 6,355 | 11,326 | 6,851 | 3,227 | 1,963 |
| **Age, in years [IQR]** | 48 [36.0, 63.0] | 47 [37.0, 59.0] | 71 [59.0, 80.0] | 85 [76.0, 90.0] | 60 [50.0, 73.0] | 86 [79.0, 91.0] | 80 [73.0, 86.0] |
| **Age, categories (%)** |  |  |  |  |  |  |  |
| 18 to 39 years | 1,437,236 (31.1) | 30,648 (31.0) | 307 (4.8) | 79 (0.7) | 665 (9.7) | 10 (0.3) | 7 (0.4) |
| 40 to 59 years | 1,785,495 (38.7) | 44,909 (45.4) | 1,355 (21.3) | 650 (5.7) | 2,665 (38.9) | 112 (3.5) | 91 (4.6) |
| 60 to 69 years | 615,198 (13.3) | 10,602 (10.7) | 1,223 (19.2) | 1,022 (9.0) | 1,304 (19.0) | 203 (6.3) | 210 (10.7) |
| 70 to 79 years | 468,286 (10.1) | 6,419 (6.5) | 1,825 (28.7) | 1,969 (17.4) | 1,316 (19.2) | 571 (17.7) | 636 (32.4) |
| 80 or older years | 312,162 (6.8) | 6,373 (6.4) | 1,645 (25.9) | 7,606 (67.2) | 901 (13.2) | 2,331 (72.2) | 1,019 (51.9) |
| **Sex, female (%)** | 2,361,230 (51.1) | 57,507 (58.1) | 2,705 (42.6) | 5,679 (50.1) | 3,067 (44.8) | 1,734 (53.7) | 769 (39.2) |
| **MEDEA Deprivation Index (%)** |  |  |  |  |  |  |  |
| Quintile 1 (least deprived) | 714,183 (15.5) | 14,873 (15.0) | 815 (12.8) | 1,272 (11.2) | 876 (12.8) | 436 (13.5) | 206 (10.5) |
| Quintile 2 | 703,921 (15.2) | 15,349 (15.5) | 936 (14.7) | 1,112 (9.8) | 1,043 (15.2) | 281 (8.7) | 192 (9.8) |
| Quintile 3 | 697,074 (15.1) | 15,564 (15.7) | 1,026 (16.1) | 1,106 (9.8) | 1,215 (17.7) | 344 (10.7) | 253 (12.9) |
| Quintile 4 | 692,844 (15.0) | 15,774 (15.9) | 1,107 (17.4) | 1,014 (9.0) | 1,205 (17.6) | 222 (6.9) | 228 (11.6) |
| Quintile 5 (most deprived) | 687,062 (14.9) | 15,134 (15.3) | 1,097 (17.3) | 896 (7.9) | 1,184 (17.3) | 204 (6.3) | 195 (9.9) |
| Rural | 832,256 (18.0) | 14,670 (14.8) | 650 (10.2) | 2,271 (20.1) | 753 (11.0) | 658 (20.4) | 206 (10.5) |
| Missing | 291,037 (6.3) | 7,587 (7.7) | 724 (11.4) | 3,655 (32.3) | 575 (8.4) | 1,082 (33.5) | 683 (34.8) |
| **Smoking status (%)** |  |  |  |  |  |  |  |
| Never smoker | 1,834,657 (39.7) | 42,353 (42.8) | 2,548 (40.1) | 3,967 (35.0) | 3,026 (44.2) | 1,220 (37.8) | 669 (34.1) |
| Former smoker | 772,875 (16.7) | 19,340 (19.5) | 2,089 (32.9) | 3,436 (30.3) | 1,901 (27.7) | 1,020 (31.6) | 730 (37.2) |
| Current smoker | 712,739 (15.4) | 13,820 (14.0) | 302 (4.8) | 839 (7.4) | 334 (4.9) | 132 (4.1) | 68 (3.5) |
| Missing | 1,298,106 (28.1) | 23,438 (23.7) | 1,416 (22.3) | 3,084 (27.2) | 1,590 (23.2) | 855 (26.5) | 496 (25.3) |
| **Comorbidities (%)** |  |  |  |  |  |  |  |
| Autoimmune condition | 259,234 (5.6) | 6,322 (6.4) | 632 (9.9) | 1,292 (11.4) | 586 (8.6) | 346 (10.7) | 244 (12.4) |
| Chronic kidney disease | 201,258 (4.4) | 4,167 (4.2) | 1,137 (17.9) | 3,708 (32.7) | 675 (9.9) | 1,076 (33.3) | 616 (31.4) |
| Chronic obstructive pulmonary disease | 119,532 (2.6) | 2,476 (2.5) | 596 (9.4) | 1,457 (12.9) | 344 (5.0) | 342 (10.6) | 271 (13.8) |
| Dementia | 42,504 (0.9) | 2,011 (2.0) | 334 (5.3) | 2,707 (23.9) | 200 (2.9) | 1,094 (33.9) | 261 (13.3) |
| Heart disease | 516,140 (11.2) | 11,076 (11.2) | 2,091 (32.9) | 5,700 (50.3) | 1,456 (21.3) | 1,470 (45.6) | 982 (50.0) |
| Hyperlipidaemia | 505,102 (10.9) | 11,015 (11.1) | 1,215 (19.1) | 1,581 (14.0) | 1,189 (17.4) | 480 (14.9) | 389 (19.8) |
| Hypertension | 687,358 (14.9) | 14,149 (14.3) | 1,947 (30.6) | 3,831 (33.8) | 1,725 (25.2) | 1,115 (34.6) | 694 (35.4) |
| Obesity | 1,144,442 (24.8) | 27,840 (28.1) | 3,197 (50.3) | 4,863 (42.9) | 3,014 (44.0) | 1,467 (45.5) | 1,031 (52.5) |
| Type 2 diabetes | 317,005 (6.9) | 6,239 (6.3) | 1,354 (21.3) | 2,608 (23.0) | 1,004 (14.7) | 741 (23.0) | 541 (27.6) |
| **Without a prior diagnosis of cancer (%)** | 4,357,710 (94.4) | 93,558 (94.5) | 5,312 (83.6) | 7,970 1(70.4) | 6,116 (89.3) | 2,631 (81.5) | 1,522 (77.5) |
| **With a prior diagnosis of cancer, by years since cancer diagnosis (%)** |  |  |  |  |  |  |  |
| ≥5 years* | 167,053 (3.6) | 3,464 (3.5) | 670 (10.5) | 1,714 (15.1) | 464 (6.8) | 379 (11.7) | 293 (14.9) |
| 1-5 years* | 72,033 (1.6) | 1,466 (1.5) | 268 (4.2) | 911 (8.0) | 211 (3.1) | 149 (4.6) | 110 (5.6) |
| <1 year* | 21,581 (0.5) | 463 (0.5) | 105 (1.7) | 731 (6.5) | 60 (0.9) | 68 (2.1) | 38 (1.9) |
| **Cancer type [ICD-10-CM code] (%)** |  |  |  |  |  |  |  |
| **Haematological** | 21,637 (0.5) | 513 (0.5) | 135 (2.1) | 268 (2.4) | 80 (1.2) | 64 (2.0) | 53 (2.7) |
| Leukaemia [C91-C95] | 5,111 (0.1) | 127 (0.1) | 24 (0.4) | 32 (0.3) | 14 (0.2) | 9 (0.3) | 6 (0.3) |
| Non-Hodgkin lymphoma [C82-C96] | 7,402 (0.2) | 175 (0.2) | 57 (0.9) | 109 (1.0) | 32 (0.5) | 37 (1.1) | 27 (1.4) |
| Hodgkin's lymphoma [C81] | 2,249 (0.0) | 48 (0.0) | 24 (0.4) | 66 (0.6) | 7 (0.1) | 7 (0.2) | 9 (0.5) |
| Multiple myeloma [C90] | 2,724 (0.1) | 60 (0.1) | 6 (0.1) | 4 (0.0) | 8 (0.1) | 0 (0.0) | 2 (0.1) |
| Other haematological [C96] | 4,151 (0.1) | 103 (0.1) | 24 (0.4) | 57 (0.5) | 19 (0.3) | 11 (0.3) | 9 (0.5) |
| **Solid** | 239,030 (5.2) | 4,880 (4.9) | 908 (14.3) | 3,088 (27.3) | 655 (9.6) | 532 (16.5) | 388 (19.8) |
| Breast [C50] | 58,611 (1.3) | 1,236 (1.2) | 148 (2.3) | 402 (3.5) | 117 (1.7) | 96 (3.0) | 46 (2.3) |
| Prostate [C61] | 37,141 (0.8) | 682 (0.7) | 186 (2.9) | 440 (3.9) | 122 (1.8) | 103 (3.2) | 87 (4.4) |
| Colorectal [C18-C21] | 36,071 (0.8) | 687 (0.7) | 170 (2.7) | 539 (4.8) | 121 (1.8) | 108 (3.3) | 82 (4.2) |
| Bladder [C67] | 20,592 (0.4) | 421 (0.4) | 132 (2.1) | 350 (3.1) | 61 (0.9) | 64 (2.0) | 56 (2.9) |
| Skin melanoma [C43] | 7,569 (0.2) | 140 (0.1) | 42 (0.7) | 314 (2.8) | 20 (0.3) | 19 (0.6) | 18 (0.9) |
| Kidney [C64] | 7,353 (0.2) | 162 (0.2) | 22 (0.3) | 64 (0.6) | 27 (0.4) | 14 (0.4) | 8 (0.4) |
| Lung [C33-C34] | 7,911 (0.2) | 159 (0.2) | 28 (0.4) | 92 (0.8) | 27 (0.4) | 17 (0.5) | 15 (0.8) |
| Corpus uterus [C54-C55] | 12,956 (0.3) | 291 (0.3) | 38 (0.6) | 74 (0.7) | 33 (0.5) | 14 (0.4) | 20 (1.0) |
| Thyroid [C73] | 6,449 (0.1) | 186 (0.2) | 12 (0.2) | 12 (0.1) | 14 (0.2) | 4 (0.1) | 4 (0.2) |
| Head and neck [C00-C14] | 5,770 (0.1) | 106 (0.1) | 21 (0.3) | 71 (0.6) | 10 (0.1) | 9 (0.3) | 8 (0.4) |
| Cervix [C53] | 3,313 (0.1) | 81 (0.1) | 6 (0.1) | 72 (0.6) | 7 (0.1) | 5 (0.2) | 4 (0.2) |
| Ovary [C56] | 3,628 (0.1) | 64 (0.1) | 10 (0.2) | 80 (0.7) | 8 (0.1) | 15 (0.5) | 3 (0.2) |
| Stomach [C16] | 3,889 (0.1) | 82 (0.1) | 6 (0.1) | 39 (0.3) | 9 (0.1) | 11 (0.3) | 5 (0.3) |
| Larynx [C32] | 3,317 (0.1) | 46 (0.0) | 18 (0.3) | 66 (0.6) | 8 (0.1) | 5 (0.2) | 8 (0.4) |
| Brain and Central Nervous System [C70-C72, C75.1-C75.3] | 2,051 (0.0) | 37 (0.0) | 8 (0.1) | 97 (0.9) | 6 (0.1) | 2 (0.1) | 3 (0.2) |
| Testis [C62] | 3,979 (0.1) | 108 (0.1) | 4 (0.1) | 25 (0.2) | 8 (0.1) | 2 (0.1) | 2 (0.1) |
| Liver [C22] | 1,622 (0.0) | 43 (0.0) | 11 (0.2) | 98 (0.9) | 7 (0.1) | 9 (0.3) | 1 (0.1) |
| Bone and cartilage [C40-C41] | 763 (0.0) | 15 (0.0) | 6 (0.1) | 41 (0.4) | 2 (0.0) | 8 (0.2) | 2 (0.1) |
| Pancreas [C25] | 479 (0.0) | 19 (0.0) | 3 (0.0) | 23 (0.2) | 4 (0.1) | 2 (0.1) | 2 (0.1) |
| Oesophagus [C15] | 2,763 (0.1) | 51 (0.1) | 0 (0.0) | 1 (0.0) | 1 (0.0) | 1 (0.0) | 0 (0.0) |
| Gallbladder [C23-C24] | 1,944 (0.0) | 43 (0.0) | 1 (0.0) | 15 (0.1) | 8 (0.1) | 2 (0.1) | 1 (0.1) |
| Other solid cancers | 10,859 (0.2%) | 221 (0.2%) | 36 (0.6%) | 173 (1.5%) | 35 (0.5%) | 22(0.7%) | 13(0.7%) |

Notes: * Years since cancer diagnosis to the index date (1 March 2020). The MEDEA deprivation index is calculated at the census tract level in urban areas. Other solid cancers include other solid cancers, cancers of unspecified site [C76, C80] and more than one cancer (i.e. patients that had more than one cancer recorded on the same date). Abbreviations: IQR, interquartile range; ICD-10-CM, International Classification for Diseases, 10th revision Clinical Modification

### Figure S4. Hazard Ratios of COVID-19 outcomes in patients with cancer compared to patients without cancer, using different adjustment strategies.

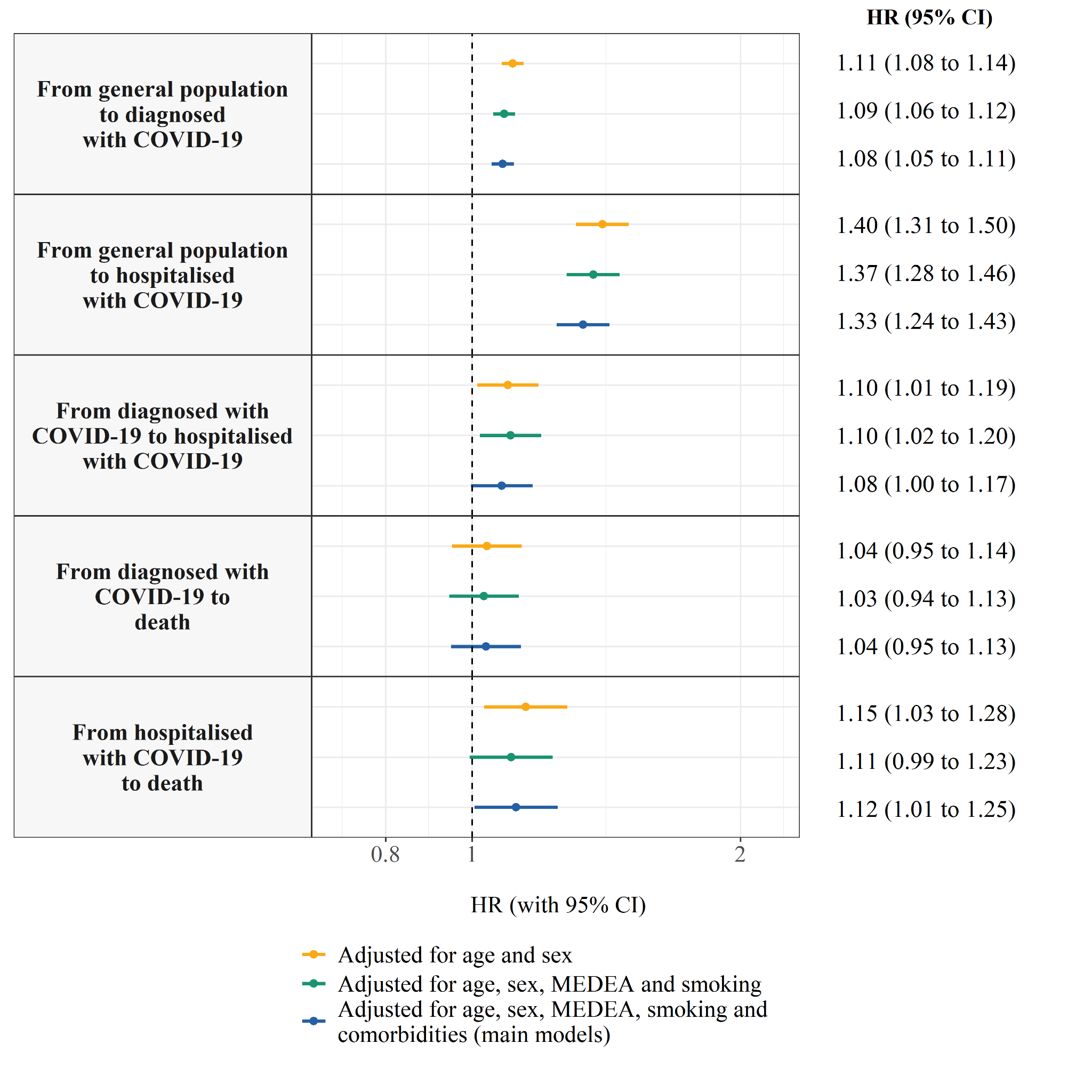

Notes: The comorbidities included are autoimmune conditions, chronic kidney disease, chronic obstructive pulmonary disease, dementia, heart disease, hyperlipidaemia, hypertension, type 2 diabetes, and obesity. Abbreviations: HR, Hazard Ratio; CI, Confidence Interval, MEDEA: MEDEA deprivation index.

### Table S2. Adjusted Hazard Ratios of COVID-19 outcomes in patients with cancer (overall and by years since cancer diagnosis) compared to patients without cancer, stratified by sex.

| **Transition** | | **Sex** | **aHR (95% CI) in patients with cancer** | | | |
| --- | --- | --- | --- | --- | --- | --- |
| **From** | **To** |  | **Overall** | **≥5 years*** | **1-5 years*** | **<1 year*** |
| General population | Diagnosed with COVID-19 | Female | 1.05 (1.01 to 1.09) | 1.08 (1.03 to 1.13) | 1.00 (0.93 to 1.08) | 0.91 (0.79 to 1.04) |
|  |  | Male | 1.08 (1.03 to 1.12) | 1.05 (1.00 to 1.11) | 1.05 (0.97 to 1.13) | 1.33 (1.17 to 1.50) |
|  | Hospitalised with COVID-19 | Female | 1.41 (1.26 to 1.58) | 1.23 (1.08 to 1.42) | 1.68 (1.38 to 2.04) | 2.24 (1.64 to 3.06) |
|  |  | Male | 1.26 (1.15 to 1.38) | 1.26 (1.13 to 1.40) | 1.13 (0.96 to 1.32) | 1.62 (1.27 to 2.07) |
| Diagnosed with COVID-19 | Hospitalised with COVID-19 | Female | 1.16 (1.03 to 1.31) | 1.10 (0.95 to 1.28) | 1.39 (1.12 to 1.72) | 0.88 (0.54 to 1.44) |
|  |  | Male | 1.05 (0.95 to 1.17) | 1.03 (0.90 to 1.17) | 1.09 (0.91 to 1.31) | 1.07 (0.79 to 1.44) |
|  | Death | Female | 0.96 (0.83 to 1.10) | 0.87 (0.74 to 1.03) | 1.12 (0.86 to 1.46) | 1.30 (0.86 to 1.97) |
|  |  | Male | 1.12 (0.99 to 1.27) | 0.98 (0.85 to 1.14) | 1.15 (0.93 to 1.43) | 2.23 (1.65 to 3.01) |
| Hospitalised with COVID-19 | Death | Female | 1.09 (0.90 to 1.30) | 0.97 (0.78 to 1.21) | 1.26 (0.90 to 1.78) | 1.73 (1.03 to 2.90) |
|  |  | Male | 1.12 (0.98 to 1.28) | 1.08 (0.93 to 1.27) | 1.12 (0.88 to 1.42) | 1.47 (0.96 to 2.23) |

Notes: * Years since cancer diagnosis to the index date (1 March 2020). Models are adjusted for age, the MEDEA deprivation index, smoking status, and comorbidities (autoimmune conditions, chronic kidney disease, chronic obstructive pulmonary disease, dementia, heart disease, hyperlipidaemia, hypertension, type 2 diabetes, and obesity). Abbreviations: aHR, adjusted Hazard Ratio; CI, Confidence Interval

### Table S3. Adjusted Hazard Ratios of COVID-19 outcomes in patients with cancer (overall and by years since cancer diagnosis) compared to patients without cancer, stratified by age.

| **Transition** | | **Age** | **aHR (95% CI) in patients with cancer** | | | |
| --- | --- | --- | --- | --- | --- | --- |
| **From** | **To** |  | **Overall** | **≥5 years*** | **1-5 years*** | **<1 year*** |
| General population | Diagnosed with COVID-19 | <70 years | 0.96 (0.93 to 1.00) | 0.97 (0.92 to 1.02) | 0.95 (0.89 to 1.01) | 0.92 (0.81 to 1.04) |
|  |  | ≥70 years | 1.12 (1.07 to 1.17) | 1.11 (1.05 to 1.16) | 1.09 (1.00 to 1.19) | 1.39 (1.21 to 1.61) |
|  | Hospitalised with COVID-19 | <70 years | 1.56 (1.37 to 1.77) | 1.48 (1.26 to 1.74) | 1.42 (1.14 to 1.77) | 2.37 (1.74 to 3.25) |
|  |  | ≥70 years | 1.21 (1.11 to 1.31) | 1.16 (1.06 to 1.28) | 1.24 (1.06 to 1.43) | 1.55 (1.21 to 1.99) |
| Diagnosed with COVID-19 | Hospitalised with COVID-19 | <70 years | 1.09 (0.97 to 1.24) | 1.03 (0.87 to 1.21) | 1.13 (0.92 to 1.40) | 1.34 (0.94 to 1.89) |
|  |  | ≥70 years | 1.07 (0.96 to 1.18) | 1.05 (0.93 to 1.19) | 1.22 (1.01 to 1.47) | 0.75 (0.51 to 1.08) |
|  | Death | <70 years | 2.97 (2.25 to 3.92) | 1.39 (0.86 to 2.24) | 3.44 (2.31 to 5.13) | 10.81 (6.83 to 17.12) |
|  |  | ≥70 years | 0.97 (0.88 to 1.06) | 0.92 (0.83 to 1.03) | 1.02 (0.85 to 1.23) | 1.29 (0.96 to 1.72) |
| Hospitalised with COVID-19 | Death | <70 years | 1.49 (1.10 to 2.01) | 1.32 (0.90 to 1.92) | 1.17 (0.65 to 2.10) | 4.58 (2.47 to 8.50) |
|  |  | ≥70 years | 1.07 (0.95 to 1.20) | 1.01 (0.89 to 1.16) | 1.16 (0.94 to 1.43) | 1.30 (0.88 to 1.90) |

Notes: * Years since cancer diagnosis to the index date (1 March 2020). Models are adjusted for sex, age, the MEDEA deprivation Index, smoking status, and comorbidities (autoimmune conditions, chronic kidney disease, chronic obstructive pulmonary disease, dementia, heart disease, hyperlipidaemia, hypertension, type 2 diabetes, and obesity). Abbreviations: aHR, adjusted Hazard Ratio; CI, Confidence Interval

### Table S4. Adjusted Hazard Ratios of COVID-19 outcomes in patients with cancer (overall and by years since cancer diagnosis) compared to patients without cancer, stratified by solid or haematological cancer.

| **Transition** | | **Cancer type,**  **number of events (overall)** | **aHR (95% CI) in patients with cancer** | | | |
| --- | --- | --- | --- | --- | --- | --- |
| **From** | **To** |  | **Overall** | **≥5 years*** | **1-5 years*** | **<1 year*** |
| General population | Diagnosed with COVID-19 | Haematological, n=513 | 1.20 (1.10 to 1.31) | 1.12 (1.00 to 1.25) | 1.23 (1.05 to 1.45) | 1.77 (1.38 to 2.28) |
|  |  | Solid, n=4,880 | 1.07 (1.04 to 1.11) | 1.09 (1.05 to 1.13) | 1.03 (0.98 to 1.09) | 1.07 (0.97 to 1.18) |
|  | Hospitalised with COVID-19 | Haematological, n=135 | 2.51 (2.12 to 2.98) | 2.01 (1.57 to 2.56) | 2.61 (1.92 to 3.53) | 6.18 (4.31 to 8.86) |
|  |  | Solid, n=908 | 1.24 (1.15 to 1.33) | 1.21 (1.11 to 1.32) | 1.21 (1.06 to 1.39) | 1.49 (1.19 to 1.87) |
| Diagnosed with COVID-19 | Hospitalised with COVID-19 | Haematological, n=80 | 1.37 (1.10 to 1.71) | 0.96 (0.68 to 1.36) | 1.85 (1.31 to 2.60) | 2.24 (1.34 to 3.76) |
|  |  | Solid, n=655 | 1.05 (0.97 to 1.14) | 1.05 (0.95 to 1.16) | 1.12 (0.96 to 1.30) | 0.85 (0.64 to 1.14) |
|  | Death | Haematological, n=64 | 1.08 (0.84 to 1.39) | 1.02 (0.72 to 1.43) | 0.89 (0.57 to 1.38) | 3.11 (1.67 to 5.81) |
|  |  | Solid, n=532 | 1.03 (0.94 to 1.13) | 0.91 (0.82 to 1.02) | 1.20 (1.00 to 1.44) | 1.69 (1.30 to 2.19) |
| Hospitalised with COVID-19 | Death | Haematological, n=53 | 1.73 (1.31 to 2.28) | 1.55 (1.04 to 2.31) | 1.92 (1.24 to 2.99) | 1.92 (0.96 to 3.86) |
|  |  | Solid, n=388 | 1.07 (0.96 to 1.20) | 1.03 (0.90 to 1.17) | 1.07 (0.86 to 1.32) | 1.56 (1.09 to 2.25) |

Notes: * Years since cancer diagnosis to the index date (1 March 2020). Models for specific cancer types include patients without cancer and patients with the cancer type of interest. Models are adjusted for sex, age, the MEDEA deprivation Index, smoking status, and comorbidities (autoimmune conditions, chronic kidney disease, chronic obstructive pulmonary disease, dementia, heart disease, hyperlipidaemia, hypertension, type 2 diabetes, and obesity). Abbreviations: aHR, adjusted Hazard Ratio; CI, Confidence Interval

### Figure S5. Adjusted Hazard Ratios of COVID-19 outcomes in patients with cancer (overall and by years since the cancer diagnosis) compared to patients without cancer, stratified by solid cancer type.

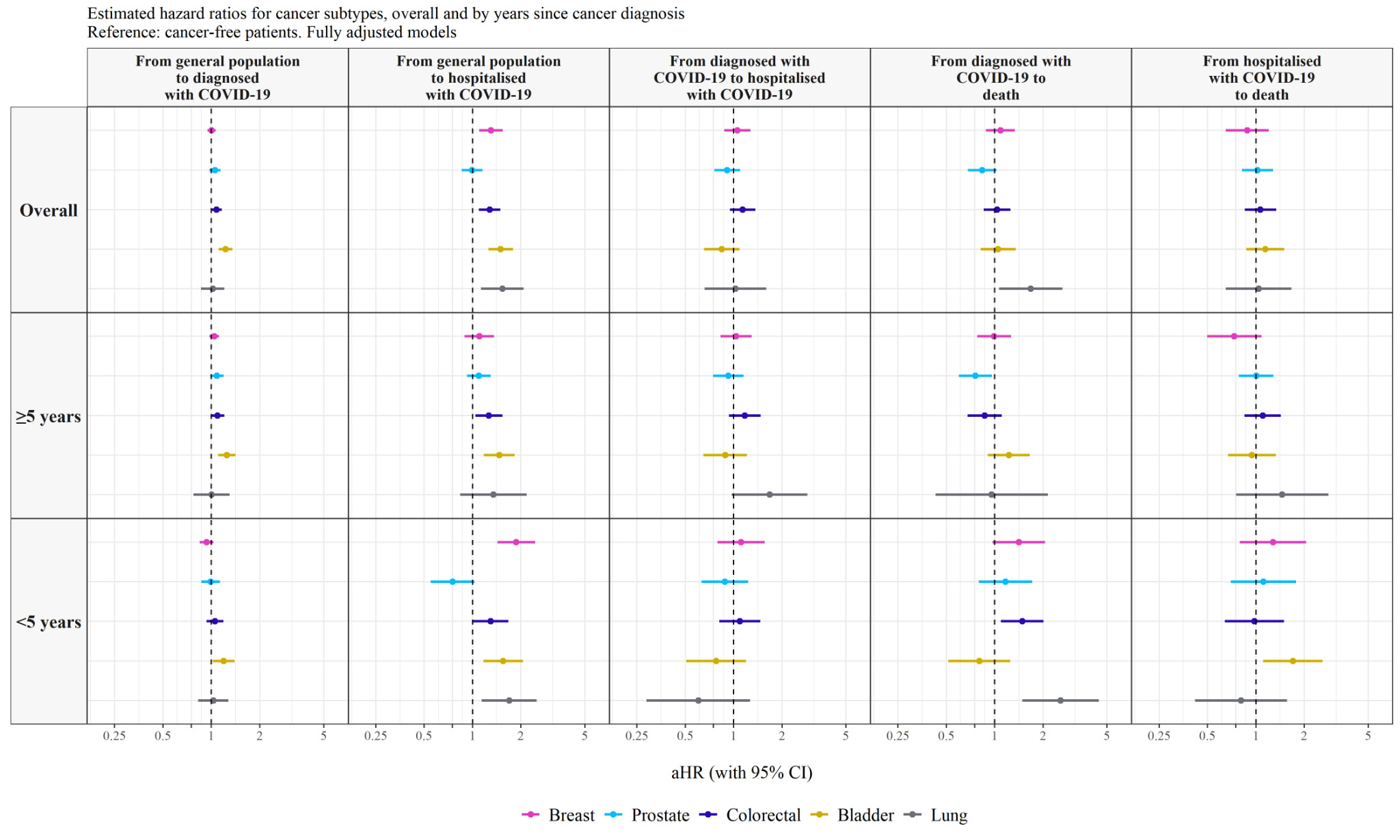

Notes: Models for specific cancer types include patients without cancer and patients with the cancer type of interest; models for prostate and breast cancer include only males and females, respectively. Models are adjusted for sex, age, smoking status, the MEDEA deprivation index, smoking status, and comorbidities (autoimmune conditions, chronic kidney disease, chronic obstructive pulmonary disease, dementia, heart disease, hyperlipidaemia, hypertension, type 2 diabetes, and obesity). Abbreviations: aHR, adjusted Hazard Ratio; CI, Confidence Interval

### Table S5. Adjusted Hazard Ratios of COVID-19 outcomes in patients with cancer (overall and by years since cancer diagnosis) compared to patients without cancer, stratified by solid cancer type.

| **Transition** | | **Solid cancer type,**  **number of events (overall)** | **aHR (95% CI) in patients with cancer** | | |
| --- | --- | --- | --- | --- | --- |
| **From** | **To** |  | **Overall** | **≥5 years*** | **<5 years*** |
| General population | Diagnosed with COVID-19 | Breast n=1,236 | 1.00 (0.95 to 1.06) | 1.04 (0.97 to 1.12) | 0.93 (0.85 to 1.03) |
|  |  | Prostate, n=682 | 1.05 (0.97 to 1.14) | 1.08 (0.99 to 1.19) | 0.99 (0.86 to 1.13) |
|  |  | Colorectal, n=687 | 1.07 (1.00 to 1.16) | 1.09 (0.99 to 1.20) | 1.05 (0.93 to 1.19) |
|  |  | Bladder, n=421 | 1.22 (1.11 to 1.35) | 1.25 (1.10 to 1.41) | 1.19 (1.02 to 1.39) |
|  |  | Lung, n= 140 | 1.02 (0.86 to 1.20) | 0.99 (0.76 to 1.28) | 1.03 (0.83 to 1.28) |
|  | Hospitalised with COVID-19 | Breast, n=148 | 1.30 (1.10 to 1.54) | 1.10 (0.89 to 1.36) | 1.87 (1.43 to 2.45) |
|  |  | Prostate, n=186 | 0.99 (0.85 to 1.16) | 1.09 (0.92 to 1.30) | 0.75 (0.55 to 1.03) |
|  |  | Colorectal, n=170 | 1.28 (1.10 to 1.49) | 1.27 (1.04 to 1.53) | 1.30 (1.01 to 1.67) |
|  |  | Bladder, n=132 | 1.50 (1.26 to 1.79) | 1.47 (1.18 to 1.83) | 1.55 (1.17 to 2.06) |
|  |  | Lung, n=42 | 1.53 (1.13 to 2.08) | 1.35 (0.84 to 2.18) | 1.69 (1.14 to 2.51) |
| Diagnosed with COVID-19 | Hospitalised with COVID-19 | Breast, n=117 | 1.06 (0.87 to 1.27) | 1.04 (0.83 to 1.29) | 1.12 (0.79 to 1.56) |
|  |  | Prostate, n=122 | 0.91 (0.76 to 1.10) | 0.93 (0.75 to 1.15) | 0.88 (0.63 to 1.23) |
|  |  | Colorectal, n=121 | 1.14 (0.95 to 1.37) | 1.17 (0.93 to 1.48) | 1.09 (0.81 to 1.47) |
|  |  | Bladder, n=61 | 0.85 (0.66 to 1.09) | 0.89 (0.65 to 1.21) | 0.78 (0.51 to 1.20) |
|  |  | Lung, n=20 | 1.03 (0.66 to 1.60) | 1.54 (0.87 to 2.70) | 0.60 (0.29 to 1.27) |
|  | Death | Breast, n=96 | 1.08 (0.88 to 1.33) | 0.99 (0.78 to 1.26) | 1.41 (0.97 to 2.05) |
|  |  | Prostate, n=103 | 0.84 (0.68 to 1.02) | 0.76 (0.60 to 0.96) | 1.17 (0.79 to 1.71) |
|  |  | Colorectal, n=108 | 1.03 (0.85 to 1.26) | 0.87 (0.68 to 1.11) | 1.48 (1.09 to 2.01) |
|  |  | Bladder, n=64 | 1.05 (0.82 to 1.35) | 1.22 (0.91 to 1.65) | 0.80 (0.51 to 1.25) |
|  |  | Lung, n=19 | 1.68 (1.06 to 2.64) | 0.96 (0.43 to 2.14) | 2.57 (1.49 to 4.46) |
| Hospitalised with COVID-19 | Death | Breast, n=46 | 0.88 (0.65 to 1.20) | 0.73 (0.50 to 1.08) | 1.28 (0.80 to 2.06) |
|  |  | Prostate, n=87 | 1.02 (0.82 to 1.28) | 1.01 (0.78 to 1.29) | 1.11 (0.70 to 1.78) |
|  |  | Colorectal, n=82 | 1.07 (0.85 to 1.34) | 1.10 (0.85 to 1.43) | 0.98 (0.64 to 1.49) |
|  |  | Bladder, n=56 | 1.14 (0.87 to 1.50) | 0.95 (0.67 to 1.33) | 1.70 (1.11 to 2.60) |
|  |  | Lung, n=18 | 1.04 (0.65 to 1.67) | 1.46 (0.75 to 2.84) | 0.81 (0.42 to 1.57) |

Notes: * Years since cancer diagnosis to the index date (1 March 2020). Models for specific cancer types include patients without cancer and patients with the cancer type of interest; models for breast and prostate cancer include only females and males, respectively. Models are adjusted for sex, age, the MEDEA deprivation index, smoking status, and comorbidities (autoimmune conditions, chronic kidney disease, chronic obstructive pulmonary disease, dementia, heart disease, hyperlipidaemia, hypertension, type 2 diabetes, and obesity). Abbreviations: aHR, adjusted Hazard Ratio; CI, Confidence Interval

### Figure S6. Log-log plots for risk of having an outpatient COVID-19 diagnosis.

A. **For age categories**

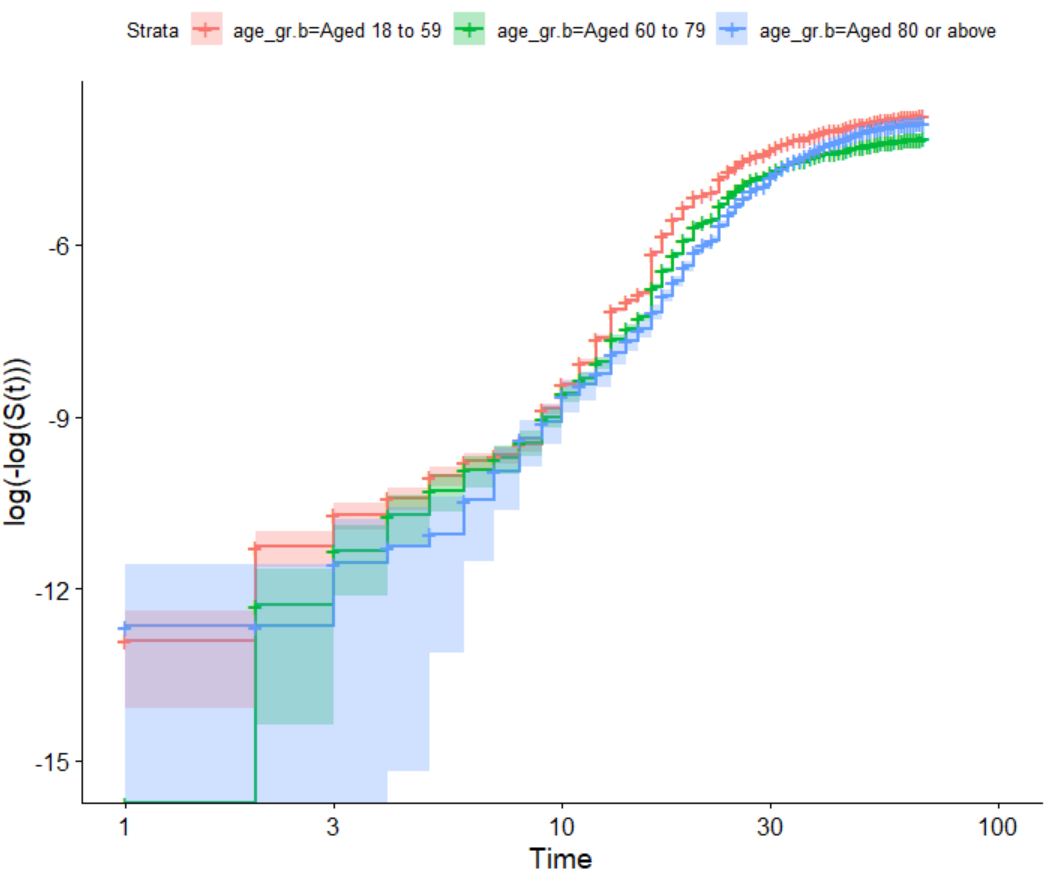

B. **For cancer status, by years since cancer diagno**sis

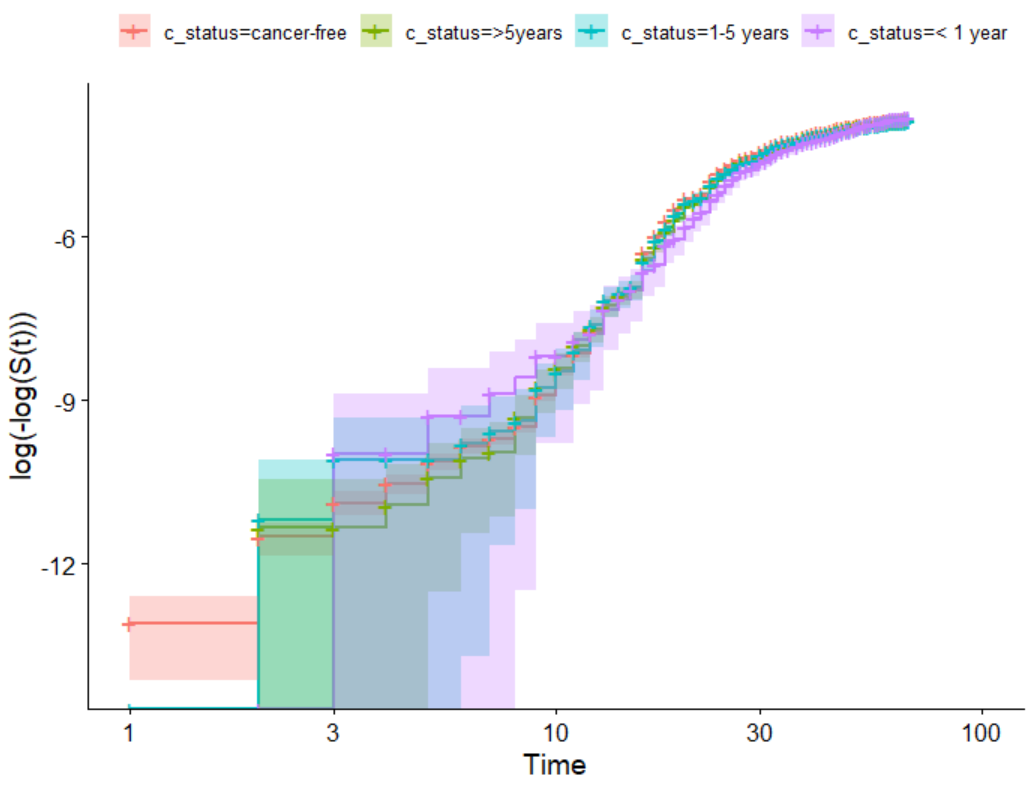

### Figure S7. Adjusted Hazard Ratios for having an outpatient COVID-19 diagnosis in patients with cancer (overall and by years since the cancer diagnosis) compared to patients without cancer, stratified by month

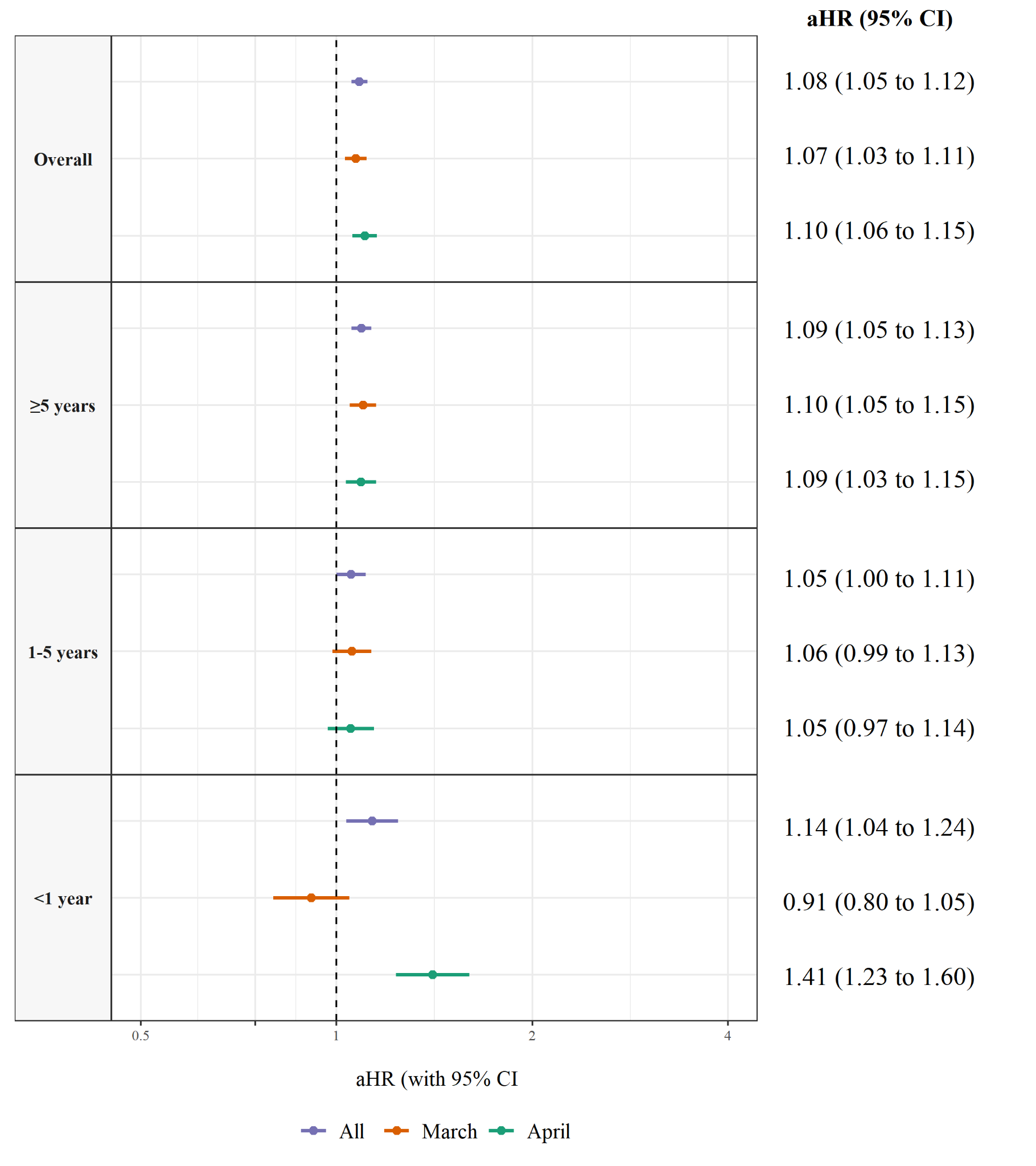

Notes: Models are adjusted for sex, age, the MEDEA deprivation Index, and comorbidities (autoimmune conditions, chronic kidney disease, chronic obstructive pulmonary disease, dementia, heart disease, hyperlipidaemia, hypertension, type 2 diabetes, and obesity). Abbreviations: aHR, adjusted Hazard Ratio; CI, Confidence Interval

### Figure S8. Adjusted Hazard Ratios of COVID-19 outcomes in patients with cancer compared to patients without cancer, by population included (all and restricting participants to never smokers)

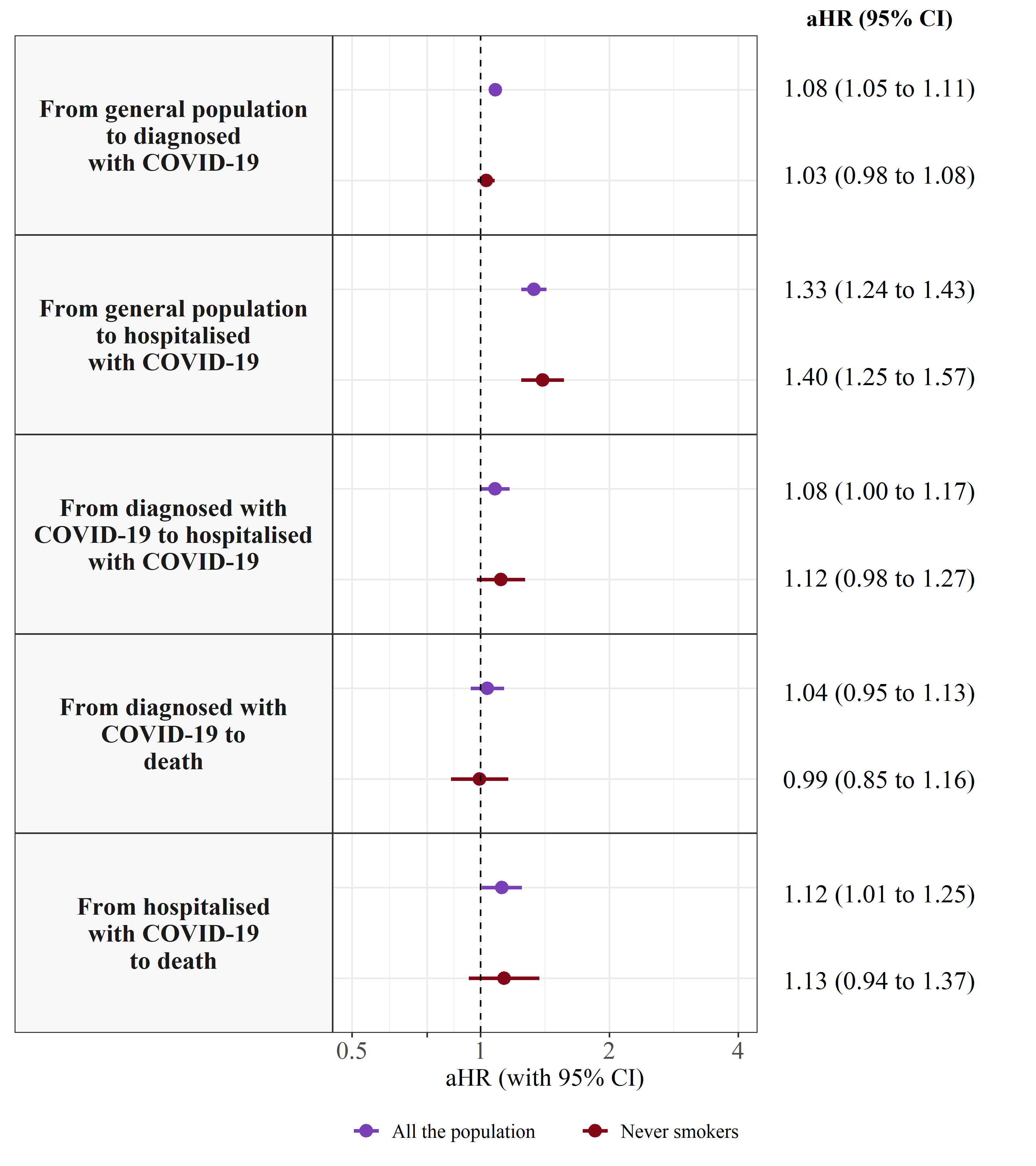

Notes: Models are adjusted for sex, age, the MEDEA deprivation Index, and comorbidities (autoimmune conditions, chronic kidney disease, chronic obstructive pulmonary disease, dementia, heart disease, hyperlipidaemia, hypertension, type 2 diabetes, and obesity). Models including all the population are also adjusted for smoking status. Abbreviations: aHR, adjusted Hazard Ratio; CI, Confidence Interval

### Table S6. Comparison of baseline characteristics among individuals with complete information on all covariates vs individuals with missing information on the MEDEA deprivation index and/or smoking status.

|  | **With complete information on covariates** | **Without MEDEA or smoking status** | **Standardized Mean Difference** |
| --- | --- | --- | --- |
| n | 3,115,935 | 1,502,442 |  |
| Age, median [IQR] | 49 [36.0, 64.0] | 47 [36.0, 61.0] | 0.063 |
| **Age, categories (%)** |  |  | 0.133 |
| 18 to 39 | 959,224 (30.8) | 478,012 (31.8) |  |
| 40 to 59 | 1,161,860 (37.3) | 623,635 (41.5) |  |
| 60 to 69 | 444,882 (14.3) | 170,316 (11.3) |  |
| 70 to 79 | 340,542 (10.9) | 127,744 (8.5) |  |
| 80 or older | 209,427 (6.7) | 102,735 (6.8) |  |
| **Sex, female (%)** | 1,639,430 (52.6) | 721,800 (48.0) | 0.092 |
| **MEDEA Deprivation Index (%)** |  |  | 0.696 |
| Quintile 1 (least deprived) | 499,651 (16.0) | 214,532 (14.3) |  |
| Quintile 2 | 501,888 (16.1) | 202,033 (13.4) |  |
| Quintile 3 | 501,880 (16.1) | 195,194 (13.0) |  |
| Quintile 4 | 501,959 (16.1) | 190,885 (12.7) |  |
| Quintile 5 (most deprived) | 501,993 (16.1) | 185,069 (12.3) |  |
| Rural | 608,564 (19.5) | 223,692 (14.9) |  |
| Missing | 0 (0.0) | 291,037 (19.4) |  |
| **Smoking status (%)** |  |  | 3.565 |
| Never smoker | 1,724,016 (55.3) | 110,641 (7.4) |  |
| Former smoker | 725,290 (23.3) | 47,585 (3.2) |  |
| Current smoker | 666,629 (21.4) | 46,110 (3.1) |  |
| Missing | 0 (0.0) | 1,298,106 (86.4) |  |
| **Comorbidities** |  |  |  |
| Autoimmune condition | 191,028 (6.1) | 68,206 (4.5) | 0.071 |
| Chronic kidney disease | 145,800 (4.7) | 55,458 (3.7) | 0.049 |
| Chronic obstructive pulmonary disease | 89,240 (2.9) | 30,292 (2.0) | 0.055 |
| Dementia | 28,197 (0.9) | 14,307 (1.0) | 0.005 |
| Heart disease | 382,684 (12.3) | 133,456 (8.9) | 0.111 |
| Hyperlipidaemia | 386,108 (12.4) | 118,994 (7.9) | 0.148 |
| Hypertension | 536,794 (17.2) | 150,564 (10.0) | 0.211 |
| Obesity | 873,059 (28.0) | 271,383 (18.1) | 0.238 |
| Type 2 diabetes | 239,653 (7.7) | 77,352 (5.1) | 0.104 |
| **Cancer status** |  |  | 0.064 |
| Without a prior diagnosis of cancer | 2,925,498 (93.9) | 1,432,212 (95.3) |  |
| With a prior diagnosis of cancer, by years since cancer diagnosis |  |  |  |
| >5 years | 121,949 (3.9) | 45,104 (3.0) |  |
| 1-5 years | 53,026 (1.7) | 19,007 (1.3) |  |
| < 1 year | 15,462 (0.5) | 6,119 (0.4) |  |
| **Age at cancer diagnosis, median [IQR]** | 61 [50.5, 70.0] | 61 [49.7, 70.9] | 0.007 |
| **Cancer type [ICD-10-CM code] (%)** |  |  |  |
| **Haematological** | 15,640 (0.5) | 5,997 (0.4) | 0.015 |
| Leukaemia [C91-C95] | 3,717 (0.1) | 1,394 (0.1) | 0.008 |
| Non-Hodgkin lymphoma [C82-C96] | 5,317 (0.2) | 2,085 (0.1) | 0.008 |
| Hodgkin's lymphoma [C81] | 1,643 (0.1) | 606 (0.0) | 0.006 |
| Multiple myeloma [C90] | 1,940 (0.1) | 784 (0.1) | 0.004 |
| Other haematological [C96] | 3,023 (0.1) | 1,128 (0.1) | 0.007 |
| **Solid** | 174,797 (5.6) | 64,233 (4.3) | 0.062 |
| Breast [C50] | 41,834 (1.3) | 16,777 (1.1) | 0.021 |
| Prostate [C61] | 28,396 (0.9) | 8,745 (0.6) | 0.038 |
| Colorectal [C18-C21] | 26,299 (0.8) | 9,772 (0.7) | 0.022 |
| Bladder [C67] | 15,570 (0.5) | 5,022 (0.3) | 0.026 |
| Melanoma [C43] | 5,804 (0.2) | 1,765 (0.1) | 0.018 |
| Kidney [C64] | 5,248 (0.2) | 2,105 (0.1) | 0.007 |
| Lung [C33-C34] | 5,749 (0.2) | 2,162 (0.1) | 0.010 |
| Corpus uterus [C54-C55] | 9,296 (0.3) | 3,660 (0.2) | 0.011 |
| Thyroid [C73] | 4,756 (0.2) | 1,693 (0.1) | 0.011 |
| Head and neck [C00-C14] | 4,136 (0.1) | 1,634 (0.1) | 0.007 |
| Cervix [C53] | 2,387 (0.1) | 926 (0.1) | 0.006 |
| Ovary [C56] | 2,623 (0.1) | 1,005 (0.1) | 0.006 |
| Stomach [C16] | 2,734 (0.1) | 1,155 (0.1) | 0.004 |
| Larynx [C32] | 2,564 (0.1) | 753 (0.1) | 0.013 |
| Brain and Central Nervous System [C70-C72, C75.1-C75.3] | 1,462 (0.0) | 589 (0.0) | 0.004 |
| Testis [C62] | 2,859 (0.1) | 1,120 (0.1) | 0.006 |
| Liver [C22] | 1,144 (0.0) | 478 (0.0) | 0.003 |
| Bone and cartilage [C40-C41] | 581 (0.0) | 182 (0.0) | 0.005 |
| Pancreas [C25] | 341 (0.0) | 138 (0.0) | 0.002 |
| Oesophagus [C15] | 1,891 (0.1) | 872 (0.1) | 0.001 |
| Gallbladder [C23-C24] | 1,441 (0.0) | 503 (0.0) | 0.006 |
| Other solid cancers | 7,682 (0.2) | 1,104 (0.02) | 0.004 |

Notes: The MEDEA deprivation index is calculated at the census tract level in urban areas. Other solid cancers include other solid cancers, cancers of unspecified site [C76, C80] and more than one cancer (i.e. patients that had more than one cancer recorded on the same date). Abbreviations: IQR, interquartile range; ICD-10-CM, International Classification for Diseases, 10^th^ revision Clinical Modification, MEDEA: MEDEA deprivation index

### Figure S9. Adjusted Hazard Ratios of COVID-19 outcomes in patients with cancer compared to patients without cancer, after multiple imputation of missing data on the MEDEA deprivation index and/or smoking status

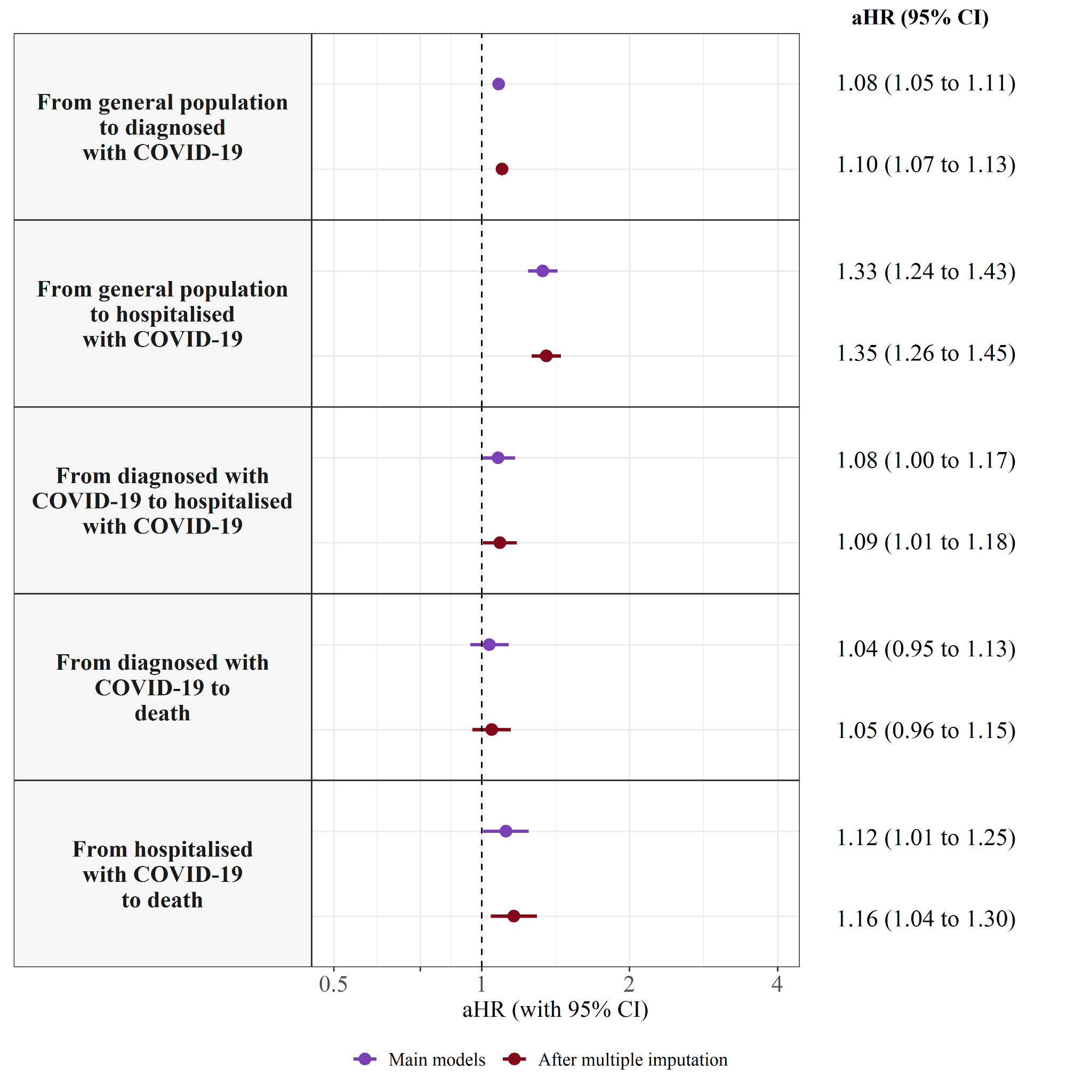

Notes: Models are adjusted for sex, age, the MEDEA deprivation Index, smoking status, and comorbidities (autoimmune conditions, chronic kidney disease, chronic obstructive pulmonary disease, dementia, heart disease, hyperlipidaemia, hypertension, type 2 diabetes, and obesity). Abbreviations: aHR, adjusted Hazard Ratio; CI, Confidence Interval
